# Genetic risk score for intracranial aneurysms to predict aneurysmal subarachnoid hemorrhage and identify associations with patient characteristics

**DOI:** 10.1101/2022.04.29.22274404

**Authors:** Mark K. Bakker, Jos P. Kanning, Gad Abraham, Amy E. Martinsen, Bendik S. Winsvold, John-Anker Zwart, Romain Bourcier, Tomonobu Sawada, Masaru Koido, Yoichiro Kamatani, Sandrine Morel, Philippe Amouyel, Stéphanie Debette, Philippe Bijlenga, Takiy Berrandou, Santhi K. Ganesh, Nabila Bouatia-Naji, Gregory Jones, Matthew Bown, HUNT All-In Stroke, CADISP group, International Consortium for Blood Pressure, International Headache Genetics Consortium, International Stroke Genetics Consortium (ISGC) Intracranial Aneurysm Working Group, Gabriël J.E. Rinkel, Jan H. Veldink, Ynte M. Ruigrok

## Abstract

**Background:** Rupture of an intracranial aneurysm (IA) causes aneurysmal subarachnoid hemorrhage (ASAH). There is no accurate prediction model for IA or ASAH in the general population. Recent discoveries in genetic risk for IA may allow improved risk prediction.

**Methods:** We constructed a genetic risk score including genetic association data for IA and 17 traits related to IA (a metaGRS) to predict ASAH incidence and IA presence. The metaGRS was trained in 1,161 IA cases and 407,392 controls in the UK Biobank and validated in combination with risk factors blood pressure, sex, and smoking in 828 IA cases and 68,568 controls from the Nordic HUNT study. We further assessed association between genetic risk load and patient characteristics in a cohort of 5,560 IA patients.

**Results:** The hazard ratio for ASAH incidence was 1.34 (95% confidence interval = 1.20-1.51) per SD increase of metaGRS. Concordance index increased from 0.63 [0.59-0.67] to 0.65 [0.62-0.69] upon including the metaGRS on top of clinical risk factors. The odds ratio for prediction of IA presence was 1.09 [95% confidence interval: 1.01-1.18], but did not improve area under the curve. The metaGRS was statistically significantly associated with age at ASAH (β=-4.82×10^−3^ per year [-6.49×10^−3^ to -3.14×10^−3^], P=1.82×10^−8^), and location at the internal carotid artery (OR=0.92 [0.86 to 0.98], P=0.0041).

**Conclusions:** The metaGRS was predictive of ASAH incidence with modest added value over clinical risk factors. Genetic risk plays a role in clinical heterogeneity of IA. Additional studies are needed to identify the biological mechanisms underlying this heterogeneity.

**KEY MESSAGES:** *What is already known on this topic:* Recent advanced in the understanding of genetic risk for IA opened and opportunity for risk prediction by combining genetic and conventional risk factors.

*What this study adds:* Here, we developed a genetic risk score based on genetic association information for IA and 17 related traits. This risk score improved prediction compared to a model including only conventional risk factors. Further, genetic risk was associated with age at ASAH and IA location.

*How this study might affect research, practice, or policy:* This study emphasizes the importance of combining conventional and genetic risk factors in prediction of IA. It provides a metric to develop an accurate risk assessment method including conventional and genetic risk factors.

## Introduction

Rupture of an intracranial aneurysm (IA) leads to aneurysmal subarachnoid hemorrhage (ASAH), a severe type of stroke causing death in one third of cases, and permanent disability in another third.^1^ It is one of few cardiovascular diseases in which women are at higher risk than men and is caused by a complex interplay of genetic factors and environmental risk factors,^2, 3^ including smoking and hypertension.^4, 5^ Aneurysmal rupture can be prevented by endovascular treatment or surgery, with relatively low risk of complications compared to the high case fatality and morbidity of ASAH.^6^ Therefore, prediction of ASAH has high potential in reducing disease burden.

Genetic risk scores (GRSs) showed potential in risk prediction of common diseases.^7^ New techniques improved prediction potential of GRSs by: 1. providing methods to include a large number of genetic variants^8^ and 2. combining GRSs for multiple traits (a so-called metaGRS), leading to improved prediction of, amongst others, coronary artery disease^9^ and ischemic stroke.^10^ These advances, combined with a substantial portion of heritability of IA being explained in the latest genome-wide association study (GWAS) of IA,^2^ provide an opportunity for genetic risk prediction of IA.

A broad spectrum of clinical heterogeneity of IAs exists, including number, size and different locations of IAs.^11, 12^ A GRS constructed with only seven single nucleotide polymorphisms (SNPs) was higher in patients with IAs at the middle cerebral artery (MCA) compared to those with IAs at other locations, in a cohort of 1,691 IA patients.^13^ In a cohort of 4,890 patients of whom 109 had an unruptured IA (UIA), a 10-SNP GRS was associated with aneurysmal diameter and volume.^14^ These studies show a potential link between genetic predisposition and patient characteristics, but additional studies using larger populations and assessing across the genome are warranted.

We created a metaGRS for IA that incorporates GWAS summary statistics for IA together with summary statistics for other stroke subtypes and risk factors for IAs, and assessed its predictive performance for ASAH incidence and IA presence. Next, we assessed how the metaGRS associates with clinical characteristics of IA patients.

## Methods

Figure 1 shows an overview of the study methods. In short, trait-level GRSs were constructed using summary statistics of the largest publicly available GWASs of IA and related traits (Supplementary Table 1). Optimal GRS model selection, and combining these GRSs into a metaGRS, was performed using the UK Biobank, a prospective cohort including 1,161 IA patients (959 with ASAH and 202 with UIA) and 407,392 controls (Figure 1A, Table 1). Predictive performance of the metaGRS was assessed in the HUNT prospective cohort study consisting of 828 IA patients (318 with ASAH and 510 with UIA) and 68,568 controls (Table 1). Associations of the metaGRS with patient characteristics were assessed in the well-phenotyped cohort of the international stroke genetics consortium (ISGC, www.strokegenetics.org) IA working group (ISGC-IA), including 5,560 IA patients of whom 3,918 with ASAH and 1,642 with UIA (Supplementary Table 2; Supplementary Data for participant selection and quality control of all cohorts).

**Table 1.** Baseline characteristics of the study populations.

| Characteristic | UK Biobank | HUNT study | ISGC-IA phenotype cohort |
| --- | --- | --- | --- |
| Age in years, mean (SD) | 65.3 (8.0) | 53.7 (17.5) | 53.3 (13.0)** |
| Women, N (%) | 220,835 (54.1%) | 36,887 (53.0%) | 3,786 (68.1%) |
| IA cases, N (%) | 1,161 (0.28%) | 828 (1.2%) | 5,560 (100%) |
| ASAH cases, N (%) | 959 (0.23%) | 318 (0.46%) | 3,916 (70.4%) |
| Controls, N | 407,392 | 68,568 | *not applicable* |
| SBP, mean (SD) | 138.2 (18.1) | 134.4 (21.1) | *no data* |
| Self-reported hypertension, N (%)*** | *no data* | *no data* | 2,150 (39.4%) |
| Ever smokers, N (%) | 123,604 (30.3%) | 35,204 (50.6%) | 3,481 (69.4%) |
| Smoking packs/day, mean (SD)* | 0.56 (0.42) | 0.39 (0.30) | *no data* |

*UK Biobank characteristics are estimated based on the most recent assessment date.*
*\*Packs/day are cumulative pack-years divided by age minus 16 based on ever smokers only.*
*\*\*In the ISGC-IA cohort only age at ASAH was recorded, and these numbers are shown.*
*\*\*\*In the ISGC-IA phenotype cohort, only self-reported hypertension was recorded.*

**Figure 1.**
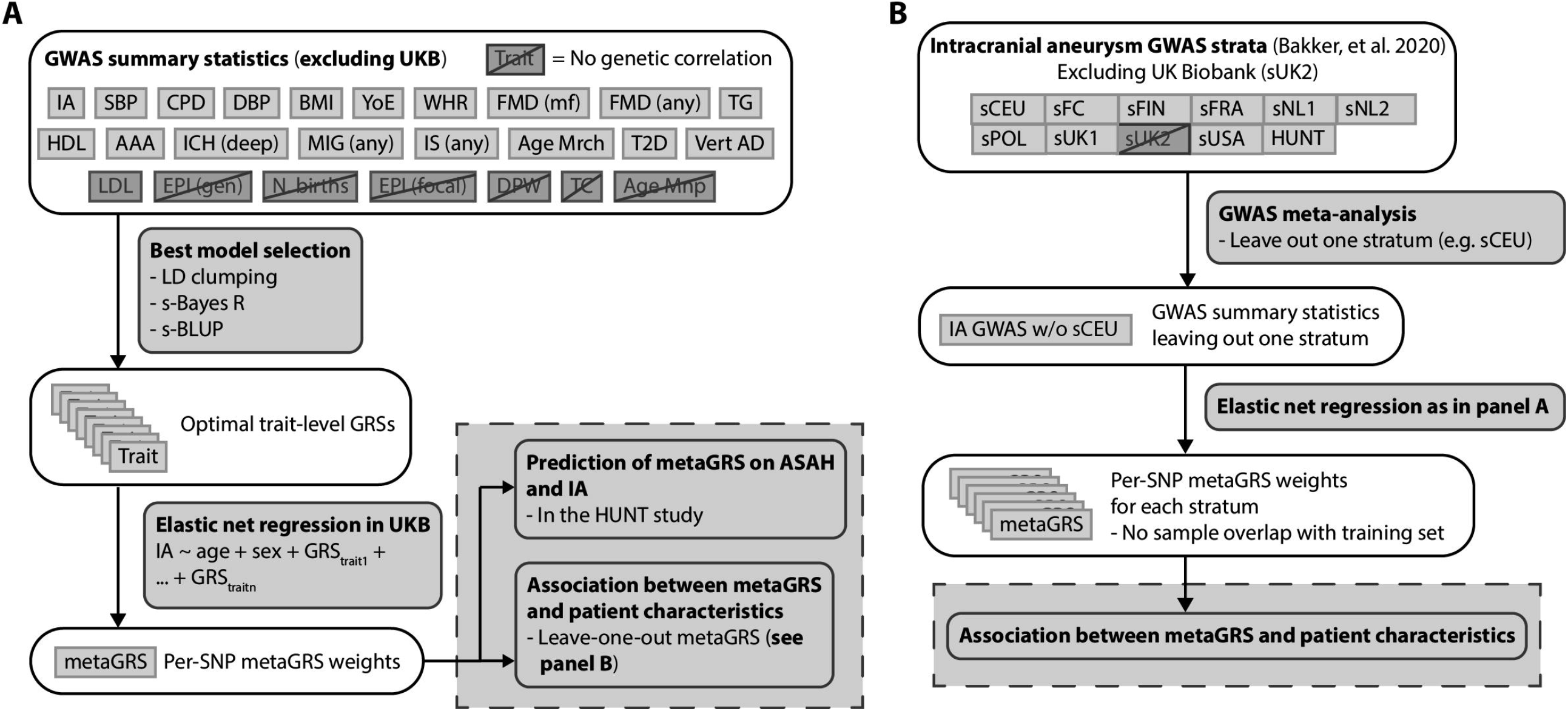
Overview of constructing the metaGRS. A) Steps to create the metaGRS used for prediction in the HUNT study. Dark grey boxes with diagonal line indicate traits without genetic correlation with IA and were not used for the next step. B) Steps to create IA GWAS summary statistics to be included in an adjusted metaGRS version for phenotype-genotype correlation analysis. Single IA GWAS strata were excluded from the IA GWAS in a leave-one-out manner prior to the ‘Best model selection’ step in panel A. IA: intracranial aneurysm, SBP: systolic blood pressure, CPD: cigarettes per day, DBP: diastolic blood pressure, BMI: body-mass index, YoE: years of education, WHR: waist-to-hip ratio, FMD (mf): multifocal fibromuscular dysplasia, FMD (any) any fibromuscular dysplasia, TG: triglycerides, HDL: high density lipoprotein, AAA: abdominal aortic aneurysm, ICH (deep): deep intracerebral hemorrhage, MIG (any): any migraine, IS (any): any ischemic stroke, Age Mrch: age at menarche, T2D: type II diabetes, Vert AD: vertebral artery dissection, LDL: low density lipoprotein, EPI (gen): generalized epilepsy, N births: number of births, EPI (focal): focal epilepsy, DPW: alcoholic drinks per week, TC: total cholesterol, Age Mnp: age at menopause, sCEU: stratum of mixed European ancestry, sFC: French Canada, sFIN: Finnish, sFRA: France, sNL1, sNL2: Netherlands, sPOL: Poland, sUK1, sUK2: United Kingdom, sUSA: United States of America, HUNT: Nordic HUNT study.

### Constructing the metaGRS

Methods of the metaGRS construction are shown in Figure 1A. Summary statistics of large GWASs of IA and 24 traits (potentially) associated with IA were obtained (Supplementary Table 1).^2^ These 24 traits included: 1. established risk factors for IA and/or ASAH, being diastolic and systolic blood pressure (SBP), smoking (cigarettes per day), and alcohol consumption (drinks per week);^4, 5, 15-19^ 2. suggestive risk factors including those related to female hormones (age at menarche, age at menopause and number of births),^20-23^ and cardiovascular disease risk (diabetes type II, body-mass index, waist-to-hip ratio, low- and high density lipoprotein levels, total cholesterol, and triglyceride levels),^15, 24-29^ migraine,^30-32^ epilepsy (focal and generalized),^2, 33^ and years of education^34, 35^ and 3. diseases genetically correlated with IA, being intracerebral hemorrhage, ischemic stroke, abdominal aortic aneurysms, and fibromuscular dysplasia (multifocal and any type).^36-39^ Only data of European-ancestry individuals were used while UK Biobank participants were excluded. Data pre-processing steps are described in the Supplementary Data.

For traits genetically correlated with IA (P<0.05) trait-level GRSs were created (Supplementary Data for more details) using three methods: LD-based clumping with 9 different LD thresholds, summary statistics-based best linear unbiased predictor^40^ and summary statistics-based BayesR^8^ (Figure 1A, Supplementary Data). To assess whether the trait-level GRSs captured risk of the respective trait, we assessed the association and predictive performance of optimal trait-level GRS with those traits in the UK Biobank. Samples with a missing genotype were ignored for association of the specific variant, while samples with missing phenotype were excluded (Supplementary Data for detailed methods).

For each trait, the trait-level GRS with highest Nagelkerke pseudo-R^2^ in predicting IA status in the UK Biobank cohort was selected (Supplementary Table 3) and subsequently jointly analyzed in an elastic-net regression to obtain per-trait weights (Figure 1A). Trait-level SNP weights were scaled according to the per-trait weights and population standard deviation, and then summed over traits to create metaGRS SNP weights (see Supplementary Data for extensive methods). These analyses were performed for the whole cohort and for men and women separately. A separate GRS was constructed only considering the IA GRS, to assess potential added value of a metaGRS over an IA-only GRS.

### Prediction of ASAH and IA by the metaGRS

Prediction by the metaGRS was evaluated in the HUNT study in two models: 1. for ASAH incidence using cox regression with age at ASAH as outcome and age at last assessment for controls as censoring time, and 2. for IA presence (including both UIA and ASAH) using a logistic regression with IA case-control status as outcome. For logistic regression, age, SBP, and average smoking packs per day since age 16 (SBP and smoking as three-knot polynomial spline), were included as covariates. For Cox regression, age was left out as covariate (further details described in the Supplementary Data). The added value of specific predictors was assessed using various models: 1. a reference model (sex and age), clinical model (only sex, age, SBP, and smoking), 2. a reference+metaGRS model, 3. a full model (clinical model + metaGRS), and 4. models leaving out a single predictor from the full model. Predictive value was determined in the HUNT study using a metaGRS created with GWAS summary statistics for IA leaving out samples from the HUNT study (Figure 1B).

### Association between metaGRS and patient characteristics

The ISGC-IA phenotype cohort of 5,560 IA patients was used to determine the association of the metaGRS with the following patient characteristics: sex, smoking status (ever or never), self-reported hypertension, age at ASAH, and family history of IA (≥1 first degree relative with ASAH and/or UIA), IA location, number of IAs (single vs multiple), rupture status (UIA vs ASAH), and aneurysmal size at rupture. Locations of IA were grouped: 1) internal carotid artery (ICA) including the ICA, ophthalmic artery, and cavernous artery, 2) posterior communicating artery (PCOM), 3) anterior cerebral arteries (ACA) including the A1 anterior segment, anterior communicating artery, and A2 segment, 4) MCA, and 5) posterior circulation (PC), including the vertebrobasilar system. IA at other locations were excluded from these analyses.

Since most cases of the ISGC-IA phenotype cohort were included in the IA GWAS, we created stratum-specific metaGRSs leaving out samples from the ISGC-IA phenotype cohort one GWAS stratum at a time from the IA GWAS summary statistics, resulting in nine metaGRS versions (Figure 1B, Supplementary Data). To control for differences in metaGRS versions between strata, we used the different cohorts of the ISGC-IA phenotype cohort (Supplementary Table 4) as covariate in all subsequent analyses.

We calculated associations between metaGRS and patient characteristics, correcting for sex and cohort using a generalized linear model. We tested whether statistically significantly associated phenotypes were independently associated from one another using a multivariate model. For each phenotype, samples with missing values were excluded for analysis of that specific phenotype. In analyses studying the association with IA location, we included only patients with one IA. Statistical analyses were done in R 4.1.2. Statistical significance was determined by Bonferroni correction for the number of phenotypes in the primary analyses: rupture status, sex, family history, IA multiplicity, five locations, age at ASAH, and size at rupture (P<0.05/11). More details on the statistics are described in the Supplementary Data.

## Results

### Constructing the metaGRS

Seventeen out of 24 traits showed genetic correlation with IA (P<0.05, Figure 1, Supplementary Table 5). Characteristics of the UK Biobank training cohort are shown in Table 1. We checked for the trait-level GRSs of which a representative phenotype was available in the UK Biobank, whether it was associates with that trait and an improvement of area under the curve (AUC) or *R*^2^ compared to a reference model (for example, the trait-level GRS for SBP being associated with SBP in the UK biobank). This was the case for all phenotypes except intracerebral hemorrhage (Supplementary Table 6). Traits with an effect in the elastic-net regression were included in the metaGRS. Elastic-net regression weights for each trait-level GRS are shown in Supplementary Table 7. In total, 7,078,955 SNPs were included in the metaGRS. In separate models trained in only men or women in the UK Biobank, 6,618,190 and 6,671,269 SNPs remained, respectively.

### Prediction of ASAH by the metaGRS

Characteristics of the HUNT study validation cohort are shown in Table 1. The metaGRS ranged from -0.83 to +0.50, with mean -0.22 and standard deviation 0.14. In the HUNT study, the metaGRS showed improved prediction of ASAH incidence compared to a reference model including only sex (hazard ratio [HR] = 1.34 [95% confidence interval, CI: 1.20 – 1.51], Supplementary Table 8). The C-index increased from 0.53 [0.49-0.56] to 0.58 [0.55-0.62] upon including the metaGRS to the reference model (Supplementary Table 9).

With a higher HR, the metaGRS seemed to outperform a GRS constructed using only summary statistics of IA (HR=1.25 [1.12-1.41], C-index=0.57 [0.53-0.61). Maximum prediction was reached upon including the metaGRS on top of clinical risk factors, where the C-index increased from 0.63 [0.59–0.67] to 0.65 [0.62–0.69] (Figure 2). Given the HR per standard deviation, HR for a person at the highest 1% of metaGRS compared to median metaGRS is 1.99 [1.52-2.61], while the highest 1% versus lowest 1% had a HR of 3.96 [2.31-6.81].

**Figure 2.**
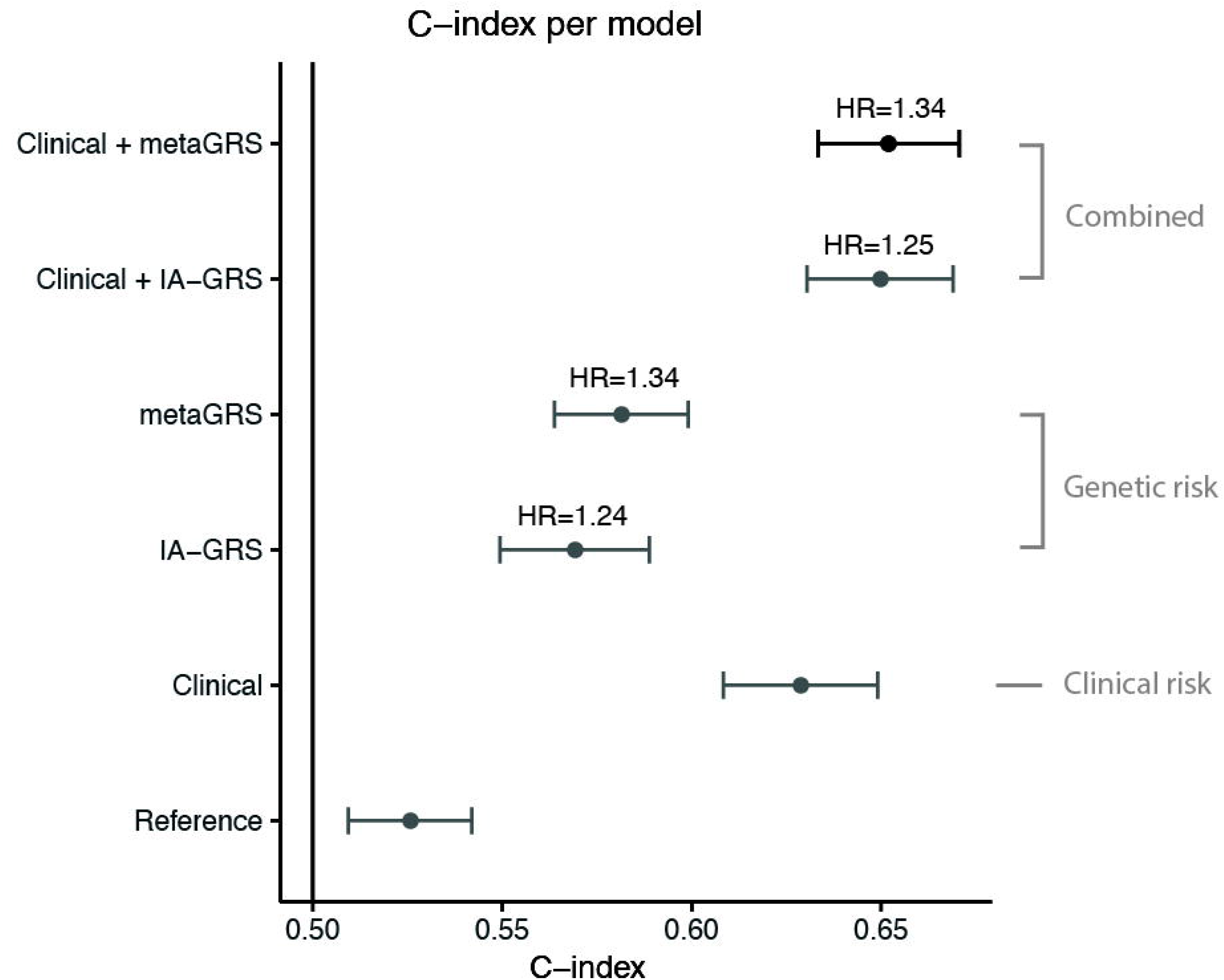
Prediction of ASAH using the metaGRS in the HUNT study. C-index according to different combinations of clinical risk factors and metaGRS are shown on the horizontal axis. Error bars denote 95% confidence intervals. HR: hazard ratio per standard deviation of the IA-only GRS or metaGRS in the model. Reference: model including only sex. Clinical: model including sex, SBP, and smoking. IA: intracranial aneurysm. GRS: genetic risk score.

In the model trained in women in the UK Biobank and validated in women in the HUNT study, the metaGRS alone had a larger effect compared to the model of both women and men and the model of men only (women: HR per SD of metaGRS=1.36 [1.18-1.60], men: 1.12 [0.93-1.34], Supplementary Figure 1, Supplementary Table 8). Similarly, clinical risk factors combined provided better prediction in women, and worse in men (women: C-index = 0.71 [0.67-0.75], men: 0.57 [0.52-0.62], Supplementary Table 9). Furthermore, metaGRS outperformed IA-only GRS in women, similar to in the whole cohort (IA-only GRS in women: HR=1.30 [1.11-1.51]).

### Prediction of IA by the metaGRS

In prediction of IA presence (either UIA or ASAH) in the HUNT study, the metaGRS provided marginal added value on top of clinical risk factors (odds ratio [OR] = 1.09 [95% CI: 1.01-1.18], Supplementary Table 10). The metaGRS did not improve prediction above a model including clinical risk factors (AUC clinical model = 0.76 [0.75–0.78], AUC clinical + metaGRS = 0.76 [0.75–0.78], Supplementary Figure 2, Supplementary Table 11). Only the predictor age showed independent added value (ΔAUC excluding age versus full model = -0.067 [-0.092 to -0.043]).

### Association between metaGRS and patient characteristics

In the ISGC-IA phenotype cohort, patients with multiple IAs had a higher metaGRS than patients with a single IA, with nominal significance (OR=1.05 [1.01–1.09], P=0.010, Table 2, Figure 3A). Younger age at ASAH was associated with a higher metaGRS (β=-4.82×10^−3^ per year [-6.49×10^−3^ to -3.14×10^−3^], P=1.82×10^−8^, Figure 3B). Accordingly, the effect of one standard deviation increase of metaGRS on age at ASAH was -1.70 [-2.30 to -1.11] years. This equates to patients with a top 5% metaGRS suffering ASAH on average 2.70 [0.65–4.74] years earlier compared to patient with a mean metaGRS, while this is 3.83 [0.95–6.70] years earlier in patients with a top versus bottom 1% metaGRS. Of all aneurysmal locations, only patients with an IA at the ICA had lower genetic risk (OR=0.92 [0.86 to 0.98], P=0.0041, Figure 3C, Supplementary Figure 3-7). This effect reduced and was not statistically significant anymore when considering ruptured IAs only (OR=0.94 [0.86–1.03], P=0.16, and Supplementary Figure 8-12). No effect was observed for sex, positive family history, rupture status of an IA, or aneurysmal size at rupture (Table 2, Supplementary Figure 13-16). A higher metaGRS was associated with hypertension (OR=1.10 [95% CI=1.06–1.14], P=3.82×10^−7^) and ever smokers (OR=1.14 [1.10–1.18], P=9.30×10^−10^, Supplementary Table 12, Supplementary Figure 17-18), which is expected due to including summary statistics for these traits in the metaGRS.

**Table 2.** Associations between metaGRS and patient characteristics in the ISGC-IA phenotype cohort.

| Subset | Phenotype | Comparison | P Value | Cases / controls | OR/Beta* | 95% CI |
| --- | --- | --- | --- | --- | --- | --- |
| All cases (N=5,560) | Ruptured | Yes / No | 0.77 | 3918 / 1642 | 1.01 | [0.96-1.05] |
|  | Sex | Men / Women | 0.74 | 1774 / 3786 | 1.01 | [0.97-1.04] |
|  | Positive Family History | Yes / No | 0.80 | 1258 / 3666 | 1.01 | [0.95-1.06] |
|  | Multiple Aneurysms | Yes / No | 0.010 | 1544 / 3955 | 1.05 | [1.01-1.09] |
| Single aneurysm all cases (N=3,820) | ICA | ICA / other | 0.0041 | 578 / 3242 | 0.92 | [0.86-0.98] |
|  | PCOM | PCOM / other | 0.94 | 517 / 3303 | 1.00 | [0.94-1.06] |
|  | ACA | ACA / other | 0.084 | 1377 / 2443 | 1.04 | [1.00-1.08] |
|  | MCA | MCA / other | 0.78 | 918 / 2902 | 1.01 | [0.96-1.06] |
|  | PC | Posterior / anterior | 0.75 | 430 / 3390 | 1.01 | [0.94-1.08] |
| Single aneurysm ASAH only (N=2,751) | ICA | ICA / other | 0.16 | 257 / 2494 | 0.94 | [0.85-1.03] |
|  | PCOM | PCOM / other | 0.93 | 445 / 2306 | 1.00 | [0.93-1.07] |
|  | ACA | ACA / other | 0.89 | 1131 / 1620 | 1.00 | [0.95-1.05] |
|  | MCA | MCA / other | 0.53 | 597 / 2154 | 1.02 | [0.96-1.08] |
|  | PC | Posterior / anterior | 0.72 | 321 / 2430 | 1.01 | [0.94-1.09] |
| ASAH only (N=3,918) | Age at Rupture | Years per SD | $1.82 \times 10^{-8}$ | N=3,893 | $-4.82 \times 10^{-3}$ | $[-6.49 \times 10^{-3} \text{ to } -3.14 \times 10^{-3}]$ |
| | Age at Rupture (outcome)** | Years per SD | $1.82 \times 10^{-8}$ | N=3,893 | -1.70 | $[-2.30 \text{ to } -1.11]$ |
| | Aneurysm Size at Rupture | Millimeter per SD | 0.35 | N=2859 | $2.61 \times 10^{-3}$ | $[-2.87 \times 10^{-3} \text{ to } 8.10 \times 10^{-3}]$ |

*\*OR/Beta: odds ratio for all phenotypes, or beta for age at rupture and aneurysm size and rupture. ISGC-IA: international stroke genetics consortium intracranial aneurysm working group. \*\*Age at rupture was also used as outcome instead of dependent variable to obtain an interpretable effect size of the association between metaGRS and age at rupture. Cases/controls: number of persons with / without for the respective phenotype, or total sample size in the case of a quantitative phenotype. CI: confidence interval. ASAH: aneurysmal subarachnoid hemorrhage. ICA: internal carotid artery. PCOM: posterior communicating artery. ACA: anterior cerebral artery. MCA: middle cerebral artery. PC: posterior circulation. per SD: effect size per standard deviation of the independent variable.*

**Figure 3.**
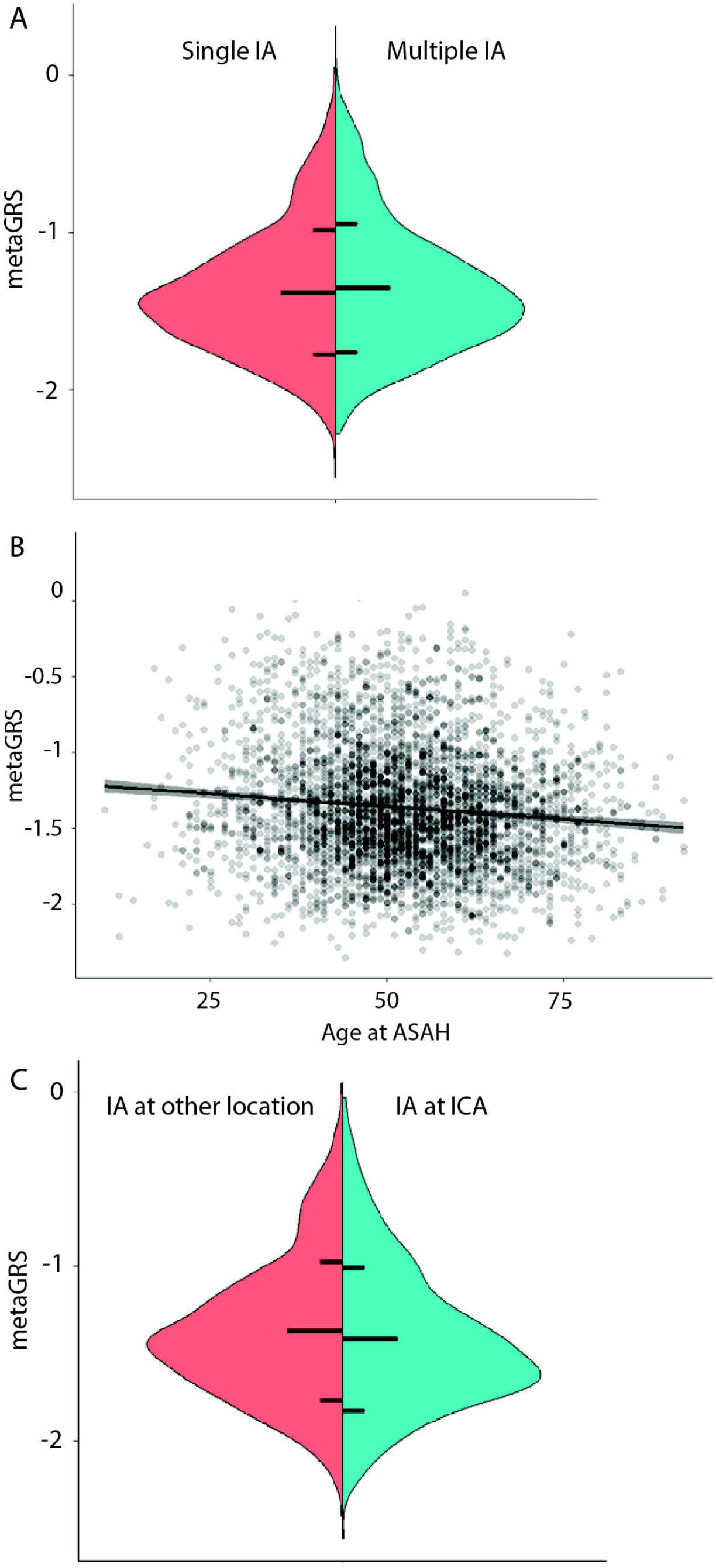
Association of metaGRS with patient characteristics. Phenotypes with at least nominally significant effect on metaGRS are shown, except hypertension and smoking status, of which proxies were included in the metaGRS. The metaGRS was transformed to mean 0, variance 1. A) metaGRS according to single or multiple IAs. Horizontal lines correspond to population mean (middle line), and mean ± one population standard deviation. B) Effect of age at ASAH on metaGRS. Line denotes regression line, and shared area is 95% confidence interval of the regression. C) Effect of IA at the ICA versus other locations, on the metaGRS. Format same as 3A. IA: intracranial aneurysm, ASAH: aneurysmal subarachnoid hemorrhage, ICA: internal carotid artery.

In the multivariate model, the association of multiple IAs with metaGRS was not independent of smoking and hypertension (OR=1.03 [0.99–1.08], P=0.16). Upon including smoking and hypertension, the effect of location at the ICA slightly reduced and became nominally significant (OR=0.93 [0.87–0.99], P=0.021), while the association between age at ASAH and metaGRS remained essentially the same (β=-5.2×10^−3^ per year [-7.03×10^−3^ to - 3.30×10^−3^], P=6.15×10^−8^) (Supplementary Table 13-15). Since the mean metaGRS was higher in persons from Finland (Supplementary Figure 19) we performed the associations analyses on multiple IAs, location at the ICA, or age at ASAH excluding these persons but the effect sizes remained essentially the same (Supplementary Table 16).

## Discussion

We created a metaGRS for IA based on GWAS summary statistics for IA and 17 IA-related traits. The metaGRS displayed added value in predicting ASAH incidence in combination with clinical risk factors, and added value compared to a GRS trained using only IA association data. Moreover, we demonstrated that prediction by the metaGRS for ASAH, which disease is seen more often in women than in men,^4, 5^ performs better in women than in men. Last we showed that the metaGRS was higher in patients who suffered ASAH at a younger age and lower in patients with an IA located at the ICA which associations were independent of hypertension and smoking.

In a previous study comparing genetic risk between 109 persons with UIA and 4,781 controls using a 10-SNP GRS no difference was found.^14^ This may be explained by the low number of patients and SNPs included in that GRS. Otherwise, it may be argued that the lack of association is caused by the fact that only UIAs were studied as in our study we were also unable to show predictive ability of the metaGRS for IA presence (combined group of UIA or ASAH). However, we think that in our study we were unable to predict IA presence because many UIAs are likely to be left undetected since UIAs are often incidental findings and therefore have a high chance of not being diagnosed in participants of observational population cohorts as used in our study.^41^ This probably resulted in low statistical power for prediction of UIA alone or in combination with ASAH. The argument that UIAs may be left undetected does not apply to the previous study on 109 persons with UIA and 4,781 controls as all participants were systematically screened with for UIAs using brain MRI.^14^

A metaGRS has been developed for other cardiovascular diseases being ischemic stroke and coronary artery disease.^9, 10^ In the present study, we found a hazard ratio per standard deviation of metaGRS for prediction of ASAH which was lower than assessed for coronary artery disease (HR=1.71, 95% CI=0.168-1.73), but higher than assessed for ischemic stroke (HR=1.26, 95% CI=1.22-1.31).^9, 10^

A previous study indicated that persons with IA at the MCA had higher genetic risk than persons with IA at other locations, while no associations were found for aneurysmal size at ASAH, patient age at ASAH, or family history of UIA/ASAH.^13, 14, 42^ We did not replicate the increased genetic load for patients with an IA at the MCA. Since this effect was found in participants from Finland and the Netherlands, and we included additional countries, this might indicate population-dependent heterogeneity. Alternatively, due to the smaller sample size (N=1,613) the previous study may have been more sensitive to false positive findings, meaning there is in no true effect. Instead, we found a decreased genetic load in patients with an IA at the ICA, which location was not analyzed in the previous study.^13^ Interestingly, IAs at the ICA also have the lowest rupture risk compared to IA at other locations.^43^ Overall, location-specific rupture risks and genetic risks followed similar patterns (highest risk for ACA, PCOM and PC, medium for MCA and lowest for ICA),^43^ albeit not statistically significant. This could mean that location-specific rupture risks are in part a downstream result of genetic risk factors, but this remains to be confirmed in future studies.

The predictive performance of the metaGRS was in part captured by the inclusion of clinical risk factors smoking and SBP. This further supports the importance of genetic predisposition for smoking and blood pressure in the risk for ASAH.^2^ Whether the remaining added value of the metaGRS is driven by additional genetic causes independent of smoking and SBP, or whether the metaGRS better captures lifelong exposure to smoking and SBP than clinical measurements of these phenotypes, remains subject for future studies.

Prevalence of IA and incidence of ASAH is higher in women than men, in contrast to most other cardiovascular diseases.^5, 41^ Here, we found improved prediction of ASAH, when the metaGRS was trained and validated in women, and reduced when trained and validated in men. Predictive value of clinical risk factors was also better in women compared to in men. Sex differences have also been described for the number and location of IAs, for which characteristics we also showed differences in genetic load.^44^ To understand the difference in genetic mechanisms of IA between men and women future investigations of genetic risk factors for IA and ASAH need to emphasize on sex differences and interactions between genetic variants and sex.

In summary, we developed a metaGRS which showed predictive ability for ASAH with modest added value over clinical risk factors, but no improved prediction of IA presence (UIA and ASAH combined). Genetic risk prediction was more effective in women, warranting further study on the sex-specific genetic causes of IA. Calibration of the metaGRS combined with clinical risk factors in independent high-risk and population cohorts are the necessary next steps before the metaGRS can be implemented for clinical use. The metaGRS was associated with age at ASAH and IA location, showing further evidence for a role of genetic risk in clinical heterogeneity of IA.

## Data Availability

All data produced are available online here: https://doi.org/10.6084/m9.figshare.19672272
(including UK Biobank for all samples, male only, and female only) and here: https://doi.org/10.6084/m9.figshare.19672269 (excluding UK Biobank for all samples, male only, and female only). Data can be used and shared upon citation of this paper and adapted if the contributions are distributed under the same conditions.

https://doi.org/10.6084/m9.figshare.19672269

## Acknowledgements

We thank the ICBP consortium, the MEGASTROKE consortium, and the ISGC for providing summary statistics. This research has been conducted using the UK Biobank Resource under application number 2532. MKB had full access to all the data in the study and takes responsibility for the integrity of the data and the accuracy of the data analysis.

The Trøndelag Health Study (HUNT) is a collaboration between HUNT Research Centre (Faculty of Medicine and Health Sciences, Norwegian University of Science and Technology NTNU), Trøndelag County Council, Central Norway Regional Health Authority, and the Norwegian Institute of Public Health. The genotyping was financed by the National Institute of health (NIH), University of Michigan, The Norwegian Research council, and Central Norway Regional Health Authority and the Faculty of Medicine and Health Sciences, Norwegian University of Science and Technology (NTNU). The genotype quality control and imputation has been conducted by the K.G. Jebsen center for genetic epidemiology, Department of public health and nursing, Faculty of medicine and health sciences, Norwegian University of Science and Technology (NTNU).

## Data availability

The metaGRS SNP weights are available under here:

https://doi.org/10.6084/m9.figshare.19672272

(including UK Biobank for all samples, male only, and female only) and here:

https://doi.org/10.6084/m9.figshare.19672269 (excluding UK Biobank for all samples, male only, and female only). Data can be used and shared upon citation of this paper and adapted if the contributions are distributed under the same conditions.

## HUNT All-In Stroke

Anne Hege Aamodt^1^, Anne Heidi Skogholt^2^, Ben M Brumpton^2^, Cristen J Willer^3^, Else C Sandset^4,5^, Espen S Kristoffersen^6,7,8^, Hanne Ellekjær^9,10^, Ingrid Heuch^8^, John-Anker Zwart^8,11,2^, Jonas B Nielsen^2,3,12^, Knut Hagen^9^, Kristian Hveem^2,13,14^, Lars G Fritsche^15^, Laurent F Thomas^2,16,17,18^, Linda M Pedersen^8^, Maiken E Gabrielsen^2^, Oddgeir L Holmen^13^, Sigrid Børte^11,2,19^, Wei Zhou^20,21^

^1^Department of Neurology, Oslo University Hospital, Oslo, Norway

^2^K. G. Jebsen Center for Genetic Epidemiology, Department of Public Health and Nursing, Faculty of Medicine and Health Sciences, Norwegian University of Science and Technology (NTNU), Trondheim, Norway

^3^Department of Internal Medicine, Division of Cardiovascular Medicine, University of Michigan, Ann Arbor, MI, 48109, USA

^4^Stroke Unit, Department of Neurology, Oslo University Hospital, Oslo, Norway

^5^Research and Development, The Norwegian Air Ambulance Foundation, Norway

^6^Department of General Practice, University of Oslo, Oslo, Norway

^7^Department of Neurology, Akershus University Hospital, Lørenskog, Norway

^8^Department of Research and Innovation, Division of Clinical Neuroscience, Oslo University Hospital, Oslo, Norway

^9^Department of Neuromedicine and Movement Science, Faculty of Medicine and Health Sciences, Norwegian University of Science and Technology (NTNU), Trondheim, Norway

^10^Stroke Unit, Department of Internal Medicine, St. Olavs Hospital, Trondheim University Hospital, Trondheim, Norway

^11^Institute of Clinical Medicine, Faculty of Medicine, University of Oslo, Oslo, Norway

^12^Department of Epidemiology Research, Statens Serum Institut, Copenhagen, Denmark

^13^HUNT Research Center, Department of Public Health and Nursing, Faculty of Medicine and Health Sciences, Norwegian University of Science and Technology (NTNU), Trondheim, Norway

^14^Department of Research, Innovation and Education, St. Olavs Hospital, Trondheim University Hospital, Trondheim, Norway

^15^Center for Statistical Genetics, Department of Biostatistics, University of Michigan, Ann Arbor, MI, 48109, USA

^16^Department of Clinical and Molecular Medicine, Norwegian University of Science and Technology (NTNU), Trondheim, Norway

^17^BioCore - Bioinformatics Core Facility, Norwegian University of Science and Technology (NTNU), Trondheim, Norway

^18^Clinic of Laboratory Medicine, St. Olavs Hospital, Trondheim University Hospital, Trondheim, Norway

^19^Research and Communication Unit for Musculoskeletal Health (FORMI), Department of Research and Innovation, Division of Clinical Neuroscience, Oslo University Hospital, Oslo, Norway

^20^Department of Computational Medicine and Bioinformatics, University of Michigan, Ann Arbor, MI, 48109, USA ^21^Analytic and Translational Genetics Unit, Massachusetts General Hospital, Boston, MA, USA

## CADISP Group

Belgium: Departments of Neurology, Erasmus University Hospital, Brussels and Laboratory of Experimental Neurology, ULB, Brussels (Shérine Abboud, Massimo Pandolfo); Department of Neurology, Leuven University Hospial (Vincent Thijs). France: Departments of Neurology, Lille University Hospital-Inserm U1171 (Didier Leys, Marie Bodenant), Sainte-Anne University Hospital, Paris (Fabien Louillet, Emmanuel Touzé, Jean-Louis Mas), Pitié-Salpêtrière University Hospital, Paris (Yves Samson, Sara Leder, Anne Léger, Sandrine Deltour, Sophie Crozier, Isabelle Méresse), Amiens University Hospital (Sandrine Canaple, Olivier Godefroy), Dijon University Hospital (Maurice Giroud, Yannick Béjot), Besançon University Hospital (Pierre Decavel, Elizabeth Medeiros, Paola Montiel, Thierry Moulin, Fabrice Vuillier); Inserm U744, Pasteur Institute, Lille (Jean Dallongeville). Finland : Department of Neurology, Helsinki University Central Hospital, Helsinki (Antti J Metso, Tiina Metso, Turgut Tatlisumak); Germany: Departments of Neurology, Heidelberg University Hospital (Caspar Grond-Ginsbach, Christoph Lichy, Manja Kloss, Inge Werner, Marie-Luise Arnold), University Hospital of Ludwigshafen (Michael Dos Santos, Armin Grau); University Hospital of München (Martin Dichgans); Department of Rehabilitation: Schmieder-Klinik, Heidelberg (Constanze Thomas-Feles, Ralf Weber, Tobias Brandt). Italy: Departments of Neurology: Brescia University Hospital (Alessandro Pezzini, Valeria De Giuli, Filomena Caria, Loris Poli, Alessandro Padovani), Milan University Hospital (Anna Bersano, Silvia Lanfranconi), University of Milano Bicocca, San Gerardo Hospital, Monza, Italy (Simone Beretta, Carlo Ferrarese), Milan Scientific Institute San Raffaele University Hospital (Giacomo Giacolone); Department of Rehabilitation, Santa Lucia Hospital, Rome (Stefano Paolucci). Switzerland: Department of Neurology, Basel University Hospital (Philippe Lyrer, Stefan Engelter, Felix Fluri, Florian Hatz, Dominique Gisler, Leo Bonati, Henrik Gensicke, Margareth Amort). UK: Clinical Neuroscience, St George’s University of London (Hugh Markus). USA : Department of Neurology, Salt Lake City, USA (Jennifer Majersik); Department of Neurology, University of Virginia, Charlottesville, USA (Bradford Worrall, Andrew Southerland); Department of Neurology, Baltimore, USA (John Cole, Steven Kittner)

## International Consortium for Blood Pressure (ICBP)

Evangelos Evangelou^1,2^, Helen R Warren^3,4^, He Gao^1,5^, Georgios Ntritsos^2^, Niki Dimou^2^, Tonu Esko^16,17^, Reedik Mägi^16^, Lili Milani^16^, Peter Almgren^18^, Thibaud Boutin^19^, Stéphanie Debette^20,21^, Jun Ding^22^, Franco Giulianini^23^, Elizabeth G Holliday^24^, Anne U Jackson^25^, Ruifang Li-Gao^26^, Wei-Yu Lin^27^, Jian’an Luan^28^, Massimo Mangino^29,30^, Christopher Oldmeadow^24^, Bram Peter Prins^31^, Yong Qian^22^, Muralidharan Sargurupremraj^21^, Nabi Shah^32,33^, Praveen Surendran^27^, Sébastien Thériault^34,35^, Niek Verweij^17,36,37^, Sara M Willems^28^, Jing-Hua Zhao^28^, Philippe Amouyel^38^, John Connell^39^, Renée de Mutsert^26^, Alex SF Doney^32^, Martin Farrall^40,41^, Cristina Menni^29^, Andrew D Morris^42^, Raymond Noordam^43^, Guillaume Paré^34^, Neil R Poulter^44^, Denis C Shields^45^, Alice Stanton^46^, Simon Thom^47^, Gonçalo Abecasis^48^, Najaf Amin^49^, Dan E Arking^50^, Kristin L Ayers^51,52^, Caterina M Barbieri^53^, Chiara Batini^54^, Joshua C Bis^55^, Tineka Blake^54^, Murielle Bochud^56^, Michael Boehnke^25^, Eric Boerwinkle^57^, Dorret I Boomsma^58^, Erwin P Bottinger^59^, Peter S Braund^60,61^, Marco Brumat^62^, Archie Campbell^63,64^, Harry Campbell^65^, Aravinda Chakravarti^50^, John C Chambers^1,5,66-68^, Ganesh Chauhan^69^, Marina Ciullo^70,71^, Massimiliano Cocca^72^, Francis Collins^73^, Heather J Cordell^51^, Gail Davies^74,75^, Martin H de Borst^76^, Eco J de Geus^58^, Ian J Deary^74,75^, Joris Deelen^77^, Fabiola Del Greco M^78^, Cumhur Yusuf Demirkale^79^, Marcus Dörr^80,81^, Georg B Ehret^50,82^, Roberto Elosua^83,84^, Stefan Enroth^85^, A Mesut Erzurumluoglu^54^, Teresa Ferreira^86,87^, Mattias Frånberg^88-90^, Oscar H Franco^91^, Ilaria Gandin^62^, Paolo Gasparini^62,72^, Vilmantas Giedraitis^92^, Christian Gieger^93-95^, Giorgia Girotto^62,72^, Anuj Goel^40,41^, Alan J Gow^74,96^, Vilmundur Gudnason^97,98^, Xiuqing Guo^99^, Ulf Gyllensten^85^, Anders Hamsten^88,89^, Tamara B Harris^100^, Sarah E Harris^63,74^, Catharina A Hartman^101^, Aki S Havulinna^102,103^, Andrew A Hicks^78^, Edith Hofer^104,105^, Albert Hofman^91,106^, Jouke-Jan Hottenga^58^, Jennifer E Huffman^19,107,108^, Shih-Jen Hwang^107,108^, Erik Ingelsson^109,110^, Alan James^111,112^, Rick Jansen^113^, Marjo-Riitta Jarvelin^1,5,114-116^, Roby Joehanes^107,117^, Åsa Johansson^85^, Andrew D Johnson^107,118^, Peter K Joshi^65^, Pekka Jousilahti^102^, J Wouter Jukema^119^, Antti Jula^102^, Mika Kähönen^120,121^, Sekar Kathiresan^17,36,122^, Bernard D Keavney^123,124^, Kay-Tee Khaw^125^, Paul Knekt^102^, Joanne Knight^126^, Ivana Kolcic^127^, Jaspal S Kooner^5,67,68,128^, Seppo Koskinen^102^, Kati Kristiansson^102^, Zoltan Kutalik^56,129^, Maris Laan^130^, Marty Larson^107^, Lenore J Launer^100^, Benjamin Lehne^1^, Terho Lehtimäki^131,132^, David CM Liewald^74,75^, Li Lin^82^, Lars Lind^133^, Cecilia M Lindgren^40,87,134^, YongMei Liu^135^, Ruth JF Loos^28,59,136^, Lorna M Lopez^74,137,138^, Yingchang Lu^59^, Leo-Pekka Lyytikäinen^131,132^, Anubha Mahajan^40^, Chrysovalanto Mamasoula^139^, Jaume Marrugat^83^, Jonathan Marten^19^, Yuri Milaneschi^140^, Anna Morgan^62^, Andrew P Morris^40,141^, Alanna C Morrison^142^, Peter J Munson^79^, Mike A Nalls^143,144^, Priyanka Nandakumar^50^, Christopher P Nelson^60,61^, Teemu Niiranen^102,145^, Ilja M Nolte^146^, Teresa Nutile^70^, Albertine J Oldehinkel^147^, Ben A Oostra^49^, Paul F O’Reilly^148^, Elin Org^16^, Sandosh Padmanabhan^64,149^, Walter Palmas^150^, Aarno Palotie^103,151,152^, Alison Pattie^75^, Brenda WJH Penninx^140^, Markus Perola^102,103,153^, Annette Peters^94,95,154^, Ozren Polasek^127,155^, Peter P Pramstaller^78,156,157^, Quang Tri Nguyen^79^, Olli T Raitakari^158,159^, Rainer Rettig^161^, Kenneth Rice^162^, Paul M Ridker^23,163^, Janina S Ried^94^, Harriëtte Riese^147^, Samuli Ripatti^103,164^, Antonietta Robino^72^, Lynda M Rose^23^, Jerome I Rotter^99^, Igor Rudan^165^, Daniela Ruggiero^70,71^, Yasaman Saba^166^, Cinzia F Sala^53^, Veikko Salomaa^102^, Nilesh J Samani^60,61^, Antti-Pekka Sarin^103^, Reinhold Schmidt^104^, Helena Schmidt^166^, Nick Shrine^54^, David Siscovick^167^, Albert V Smith^97,98^, Harold Snieder^146^, Siim Sõber^130^, Rossella Sorice^70^, John M Starr^74,168^, David J Stott^169^, David P Strachan^170^, Rona J Strawbridge^88,89^, Johan Sundström^133^, Morris A Swertz^171^, Kent D Taylor^99^, Alexander Teumer^81,172^, Martin D Tobin^54^, Maciej Tomaszewski^123,124^, Daniela Toniolo^53^, Michela Traglia^53^, Stella Trompet^119,173^, Jaakko Tuomilehto^174-177^, Christophe Tzourio^21^, André G Uitterlinden^91,178^, Ahmad Vaez^146,179^, Peter J van der Most^146^, Cornelia M van Duijn^49^, Germaine C Verwoert^91^, Veronique Vitart^19^, Uwe Völker^81,180^, Peter Vollenweider^181^, Dragana Vuckovic^62,182^, Hugh Watkins^40,41^, Sarah H Wild^183^, Gonneke Willemsen^58^, James F Wilson^19,65^, Alan F Wright^19^, Jie Yao^99^, Tatijana Zemunik^184^, Weihua Zhang^1,67^, John R Attia^24^, Adam S Butterworth^27,185^, Daniel I Chasman^23,163^, David Conen^186,187^, Francesco Cucca^188,189^, John Danesh^27,185^, Caroline Hayward^19^, Joanna MM Howson^27^, Markku Laakso^190^, Edward G Lakatta^191^, Claudia Langenberg^28^, Olle Melander^18^, Dennis O Mook-Kanamori^26,192^, Colin NA Palmer^32^, Lorenz Risch^193-195^, Robert A Scott^28^, Rodney J Scott^24^, Peter Sever^128^, Tim D Spector^29^, Pim van der Harst^196^, Nicholas J Wareham^28^, Eleftheria Zeggini^31^, Daniel Levy^107,118^, Patricia B Munroe^3,4^, Christopher Newton-Cheh^134,197,198^, Morris J Brown^3,4^, Andres Metspalu^16^, Bruce M. Psaty^201,202^, Louise V Wain^54^, Paul Elliott^1,5,203-205^, Mark J Caulfield^3,4^

1. Department of Epidemiology and Biostatistics, Imperial College London, London, UK.
2. Department of Hygiene and Epidemiology, University of Ioannina Medical School, Ioannina, Greece.
3. William Harvey Research Institute, Barts and The London School of Medicine and Dentistry, Queen Mary University of London, London, UK.
4. National Institute for Health Research, Barts Cardiovascular Biomedical Research Center, Queen Mary University of London, London, UK.
5. MRC-PHE Centre for Environment and Health, Imperial College London, London, UK.
7. Division of Epidemiology, Department of Medicine, Institute for Medicine and Public Health, Vanderbilt Genetics Institute, Vanderbilt University Medical Center, Tennessee Valley Healthcare System (626)/Vanderbilt University, Nashville, TN, USA.
8. Vanderbilt Genetics Institute, Vanderbilt Epidemiology Center, Department of Obstetrics and Gynecology, Vanderbilt University Medical Center; Tennessee Valley Health Systems VA, Nashville, TN, USA.
9. Department of Epidemiology, Emory University Rollins School of Public Health, Atlanta, GA, USA.
10. Department of Biomedical Informatics, Emory University School of Medicine, Atlanta, GA, USA.
11. Massachusetts Veterans Epidemiology Research and Information Center (MAVERIC), VA Boston Healthcare System, Boston, USA.
12. Division of Aging, Department of Medicine, Brigham and Women’s Hospital, Boston, MA, Department of Medicine, Harvard Medical School, Boston, MA, USA.
13. Atlanta VAMC and Emory Clinical Cardiovascular Research Institute, Atlanta, GA, USA.
14. VA Palo Alto Health Care System; Division of Cardiovascular Medicine, Stanford University School of Medicine, CA, USA.
15. Nephrology Section, Memphis VA Medical Center and University of Tennessee Health Science Center, Memphis, TN, USA.
16. Estonian Genome Center, University of Tartu, Tartu, Estonia.
17. Program in Medical and Population Genetics, Broad Institute of Harvard and MIT, Cambridge, MA, USA.
18. Department Clinical Sciences, Malmö, Lund University, Malmö, Sweden.
19. MRC Human Genetics Unit, MRC Institute of Genetics and Molecular Medicine, University of Edinburgh, Western General Hospital, Edinburgh, Scotland, UK
20. Department of Neurology, Bordeaux University Hospital, Bordeaux, France.
21. Univ. Bordeaux, Inserm, Bordeaux Population Health Research Center, CHU Bordeaux, Bordeaux, France.
22. Laboratory of Genetics and Genomics, NIA/NIH, Baltimore, MD, USA.
23. Division of Preventive Medicine, Brigham and Women’s Hospital, Boston, MA, USA.
24. Hunter Medical Reseach Institute and Faculty of Health, University of Newcastle, New Lambton Heights, New South Wales, Australia.
25. Department of Biostatistics and Center for Statistical Genetics, University of Michigan, Ann Arbor, MI, USA.
26. Department of Clinical Epidemiology, Leiden University Medical Center, Leiden, the Netherlands.
27. MRC/BHF Cardiovascular Epidemiology Unit, Department of Public Health and Primary Care, University of Cambridge, Cambridge, UK.
28. MRC Epidemiology Unit, University of Cambridge School of Clinical Medicine, Cambridge, UK.
29. Department of Twin Research and Genetic Epidemiology, Kings College London, London, UK.
30. NIHR Biomedical Research Centre at Guy’s and St Thomas’ Foundation Trust, London, UK.
31. Wellcome Trust Sanger Institute, Hinxton, UK.
32. Division of Molecular and Clinical Medicine, School of Medicine, University of Dundee, UK.
33. Department of Pharmacy, COMSATS Institute of Information Technology, Abbottabad, Pakistan.
34. Department of Pathology and Molecular Medicine, McMaster University, Hamilton, Canada.
35. Institut universitaire de cardiologie et de pneumologie de Québec-Université Laval,, Quebec City, Canada.
36. Cardiovascular Research Center and Center for Human Genetic Research, Massachusetts General Hospital, Boston, Massachusetts, MA, USA.
37. University of Groningen, University Medical Center Groningen, Department of Cardiology, Groningen, The Netherlands.
38. University of Lille, Inserm, Centre Hosp. Univ Lille, Institut Pasteur de Lille, UMR1167 - RID-AGE - Risk factors and molecular determinants of aging-related diseases, Epidemiology and Public Health Department, Lille, France.
39. University of Dundee, Ninewells Hospital & Medical School, Dundee,, UK.
40. Wellcome Trust Centre for Human Genetics, University of Oxford, Oxford, UK.
41. Division of Cardiovascular Medicine, Radcliffe Department of Medicine, University of Oxford, Oxford, UK.
42. Usher Institute of Population Health Sciences and Informatics, University of Edinburgh, UK.
43. Department of Internal Medicine, Section Gerontology and Geriatrics, Leiden University Medical Center, Leiden, The Netherlands.
44. Imperial Clinical Trials Unit, Stadium House, 68 Wood Lane, London, UK.
45. School of Medicine, University College Dublin, Ireland.
46. Molecular and Cellular Therapeutics, Royal College of Surgeons in Ireland, Dublin, Ireland.
47. International Centre for Circulatory Health, Imperial College London, London, UK.
48. Center for Statistical Genetics, Dept. of Biostatistics, SPH II, Washington Heights, Ann Arbor, MI, USA.
49. Genetic Epidemiology Unit, Department of Epidemiology, Erasmus MC, Rotterdam, the Netherlands.
50. Center for Complex Disease Genomics, McKusick-Nathans Institute of Genetic Medicine, Johns Hopkins University School of Medicine, Baltimore, MD, USA.
51. Institute of Genetic Medicine, Newcastle University, Newcastle upon Tyne, UK.
52. Sema4, a Mount Sinai venture, Stamford, CT, USA.
53. Division of Genetics and Cell Biology, San Raffaele Scientific Institute, Milano, Italy.
54. Department of Health Sciences, University of Leicester, Leicester, UK.
55. Cardiovascular Health Research Unit, Department of Medicine, University of Washington, Seattle, WA, USA.
56. Institute of Social and Preventive Medicine, University Hospital of Lausanne, Lausanne, Switzerland.
57. Human Genetics Center, School of Public Health, The University of Texas Health Science Center at Houston and Human Genome Sequencing Center, Baylor College of Medicine, One Baylor Plaza, Houston, TX, USA.
58. Department of Biological Psychology, Vrije Universiteit Amsterdam, EMGO+ institute, VU University medical center, Amsterdam, the Netherlands.
59. The Charles Bronfman Institute for Personalized Medicine, Icachn School of Medicine at Mount Sinai, NY, USA.
60. Department of Cardiovascular Sciences, University of Leicester, Leicester, UK.
61. NIHR Leicester Biomedical Research Centre, Glenfield Hospital, Groby Road, Leicester, UK.
62. Department of Medical, Surgical and Health Sciences, University of Trieste,, Trieste, Italy.
63. Medical Genetics Section, Centre for Genomic and Experimental Medicine, Institute of Genetics and Molecular Medicine, University of Edinburgh, Edinburgh, UK.
64. Generation Scotland, Centre for Genomic and Experimental Medicine, University of Edinburgh, Edinburgh, UK.
65. Centre for Global Health Research, Usher Institute of Population Health Sciences and Informatics, University of Edinburgh, Edinburgh, Scotland, UK
66. Lee Kong Chian School of Medicine, Nanyang Technological University, Singapore, Singapore.
67. Department of Cardiology, Ealing Hospital, Middlesex, UK.
68. Imperial College Healthcare NHS Trust, London, UK.
69. Centre for Brain Research, Indian Institute of Science, Bangalore, India.
70. Institute of Genetics and Biophysics “A. Buzzati-Traverso”, CNR, Napoli, Italy.
71. IRCCS Neuromed, Pozzilli, Isernia, Italy.
72. Institute for Maternal and Child Health IRCCS Burlo Garofolo, Trieste, Italy.
73. Medical Genomics and Metabolic Genetics Branch, National Human Genome Research Institute, NIH, Bethesda, MD, USA.
74. Centre for Cognitive Ageing and Cognitive Epidemiology, University of Edinburgh, 7 George Square, Edinburgh, UK.
75. Department of Psychology, University of Edinburgh, 7 George Square, Edinburgh, UK.
76. Department of Internal Medicine, Division of Nephrology, University of Groningen, University Medical Center Groningen, Groningen, The Netherlands.
77. Department of Molecular Epidemiology, Leiden University Medical Center, Leiden, the Netherlands.
78. Institute for Biomedicine, Eurac Research, Bolzano, Italy - Affiliated Institute of the University of Lübeck, Lübeck, Germany.
79. Mathematical and Statistical Computing Laboratory, Office of Intramural Research, Center for Information Technology, National Institutes of Health, Bethesda, MD, USA.
80. Department of Internal Medicine B, University Medicine Greifswald, Greifswald, Germany.
81. DZHK (German Centre for Cardiovascular Research), partner site Greifswald, Greifswald, Germany.
82. Cardiology, Department of Medicine, Geneva University Hospital, Geneva, Switzerland.
83. CIBERCV & Cardiovascular Epidemiology and Genetics, IMIM. Dr Aiguader
84. Barcelona, Spain. 84. Faculty of Medicine, Universitat de Vic-Central de Catalunya, Vic, Spain.
85. Department of Immunology, Genetics and Pathology, Uppsala Universitet, Science for Life Laboratory, Uppsala, Sweden.
86. Wellcome Centre for Human Genetics, University of Oxford, Roosevelt Drive, Oxford, UK.
87. Big Data Institute, Li Ka Shing Center for Health for Health Information and Discovery, Oxford University, Old Road, Oxford, UK.
88. Cardiovascular Medicine Unit, Department of Medicine Solna, Karolinska Institutet, Stockholm, Sweden.
89. Centre for Molecular Medicine, L8:03, Karolinska Universitetsjukhuset, Solna, Sweden.
90. Department of Numerical Analysis and Computer Science, Stockholm University, Stockholm, Sweden.
91. Department of Epidemiology, Erasmus MC, Rotterdam, the Netherlands.
92. Department of Public Health and Caring Sciences, Geriatrics, Uppsala, Sweden.
93. Research Unit of Molecular Epidemiology, Helmholtz Zentrum München, German Research Center for Environmental Health, Neuherberg, Germany.
94. Institute of Epidemiology, Helmholtz Zentrum München, German Research Center for Environmental Health, Neuherberg, Germany.
95. German Center for Diabetes Research (DZD e.V.), Neuherberg, Germany.
96. Department of Psychology, School of Social Sciences, Heriot-Watt University, Edinburgh, UK.
97. Faculty of Medicine, University of Iceland, Reykjavik, Iceland.
98. Icelandic Heart Association, Kopavogur, Iceland.
99. The Institute for Translational Genomics and Population Sciences, Department of Pediatrics, LABioMed at Harbor-UCLA Medical Center, Torrance, CA, USA.
100. Intramural Research Program, Laboratory of Epidemiology, Demography, and Biometry, National Institute on Aging, Bethesda, MD, USA.
101. Department of Psychiatry, University of Groningen, University Medical Center Groningen, Groningen, The Netherlands.
102. Department of Public Health Solutions, National Institute for Health and Welfare (THL), Helsinki, Finland.
103. Institute for Molecular Medicine Finland (FIMM), University of Helsinki, Helsinki, Finland.
104. Clinical Division of Neurogeriatrics, Department of Neurology, Medical University of Graz, Graz, Austria.
105. Institute for Medical Informatics, Statistics and Documentation, Medical University of Graz, Graz, Austria.
106. Department of Epidemiology, Harvard T.H. Chan School of Public Health, Boston, MA, USA.
107. National Heart, Lung and Blood Institute’s Framingham Heart Study, Framingham, MA, USA.
108. The Population Science Branch, Division of Intramural Research, National Heart Lung and Blood Institute national Institute of Health, Bethesda, MD, USA.
109. Department of Medical Sciences, Molecular Epidemiology and Science for Life Laboratory, Uppsala University, Uppsala, Sweden.
110. Division of Cardiovascular Medicine, Department of Medicine, Stanford University School of Medicine, Stanford, CA USA.
111. Department of Pulmonary Physiology and Sleep, Sir Charles Gairdner Hospital, Hospital Avenue, Nedlands, Australia.
112. School of Medicine and Pharmacology, University of Western Australia.
113. Department of Psychiatry, VU University Medical Center, Amsterdam Neuroscience, Amsterdam, the Netherlands.
114. Biocenter Oulu, University of Oulu, Oulu, Finland.
115. Center For Life-course Health Research, University of Oulu, Oulu Finland.
116. Unit of Primary Care, Oulu University Hospital, Oulu, Oulu, Finland.
117. Hebrew SeniorLife, Harvard Medical School, Boston, MA, USA.
118. Population Sciences Branch, National Heart, Lung and Blood Institute, National Institutes of Health, Bethesda, MD, USA.
119. Department of Cardiology, Leiden University Medical Center, Leiden, the Netherlands.
120. Department of Clinical Physiology, Tampere University Hospital, Tampere, Finland.
121. Department of Clinical Physiology, Finnish Cardiovascular Research Center - Tampere, Faculty of Medicine and Life Sciences, University of Tampere, Tampere, Finland.
122. Broad Institute of the Massachusetts Institute of Technology and Harvard University, Cambridge, MA, USA.
123. Division of Cardiovascular Sciences, Faculty of Biology, Medicine and Health, The University of Manchester, Manchester, UK.
124. Division of Medicine, Manchester University NHS Foundation Trust, Manchester Academic Health Science Centre, Manchester, UK
125. Department of Public Health and Primary Care, Institute of Public Health, University of Cambridge, Cambridge, UK.
126. Data Science Institute and Lancaster Medical School, Lancaster, UK.
127. Department of Public Health, Faculty of Medicine, University of Split, Croatia.
128. National Heart and Lung Institute, Imperial College London, London, UK.
129. Swiss Institute of Bioinformatics, Lausanne, Switzerland.
130. Institute of Biomedicine and Translational Medicine, University of Tartu, Tartu, Estonia.
131. Department of Clinical Chemistry, Fimlab Laboratories, Tampere, Finland.
132. Department of Clinical Chemistry, Finnish Cardiovascular Research Center - Tampere, Faculty of Medicine and Life Sciences, University of Tampere, Tampere, Finland
133. Department of Medical Sciences, Cardiovascular Epidemiology, Uppsala University, Uppsala, Sweden.
134. Program in Medical and Population Genetics, Broad Institute, Cambridge, MA, USA.
135. Division of Public Health Sciences, Wake Forest School of Medicine, Winston-Salem, NC, USA.
136. Mindich Child health Development Institute, The Icahn School of Medicine at Mount Sinai, New York, NY, USA.
137. Department of Psychiatry, Royal College of Surgeons in Ireland, Education and Research Centre, Beaumont Hospital, Dublin, Ireland.
138. University College Dublin, UCD Conway Institute, Centre for Proteome Research, UCD, Belfield, Dublin, Ireland.
139. Institute of Health and Society, Newcastle University, Newcastle upon Tyne, UK.
140. Department of Psychiatry, Amsterdam Public Health and Amsterdam Neuroscience, VU University Medical Center/GGZ inGeest, Amsterdam, The Netherlands.
141. Department of Biostatistics, University of Liverpool, Block F, Waterhouse Building, Liverpool, UK.
142. Department of Epidemiology, Human Genetics and Environmental Sciences, School of Public Health, University of Texas Health Science Center at Houston, Houston, TX, USA.
143. Data Tecnica International, Glen Echo, MD, USA.
144. Laboratory of Neurogenetics, National Institute on Aging, Bethesda, USA.
145. Department of Medicine, Turku University Hospital and University of Turku, Finland.
146. Department of Epidemiology, University of Groningen, University Medical Center Groningen, Groningen, The Netherlands.
147. Interdisciplinary Center Psychopathology and Emotion regulation (ICPE), University of Groningen, University Medical Center Groningen, Groningen, The Netherlands.
148. SGDP Centre, Institute of Psychiatry, Psychology and Neuroscience, King’s College London, London, UK.
149. British Heart Foundation Glasgow Cardiovascular Research Centre, Institute of Cardiovascular and Medical Sciences, College of Medical, Veterinary and Life Sciences, University of Glasgow, Glasgow, UK.
150. Department of Medicine, Columbia University Medical Center, New York, NY, USA.
151. Analytic and Translational Genetics Unit, Department of Medicine, Department of Neurology and Department of Psychiatry Massachusetts General Hospital, Boston, MA, USA.
152. The Stanley Center for Psychiatric Research and Program in Medical and Population Genetics, The Broad Institute of MIT and Harvard, Cambridge, MA, USA.
153. University of Tartu, Tartu, Estonia.
154. German Center for Cardiovascular Disease Research (DZHK), partner site Munich, Neuherberg, Germany.
155. Psychiatric hospital “Sveti Ivan”, Zagreb, Croatia.
156. Department of Neurology, General Central Hospital, Bolzano, Italy.
157. Department of Neurology, University of Lübeck, Lübeck, Germany.
158. Department of Clinical Physiology and Nuclear Medicine, Turku University Hospital, Turku, Finland.
159. Research Centre of Applied and Preventive Cardiovascular Medicine, University of Turku, Turku, Finland.
161. Institute of Physiology, University Medicine Greifswald, Karlsburg, Germany.
162. Department of Biostatistics University of Washington, Seattle, WA, USA.
163. Harvard Medical School, Boston MA.
164. Public health, Faculty of Medicine, University of Helsinki, Finland
165. Centre for Global Health Research, Usher Institute of Population Health Sciences and Informatics, University of Edinburgh, Scotland, UK.
166. Gottfried Schatz Research Center for Cell Signaling, Metabolism & Aging, Molecular Biology and Biochemistry, Medical University of Graz, Graz, Austria.
167. The New York Academy of Medicine, New York, NY, USA.
168. Alzheimer Scotland Dementia Research Centre, University of Edinburgh, Edinburgh, UK.
169. Institute of Cardiovascular and Medical Sciences, Faculty of Medicine, University of Glasgow, United Kingdom.
170. Population Health Research Institute, St George’s, University of London, London, UK.
171. Department of Genetics, University of Groningen, University Medical Center Groningen, Groningen, The Netherlands.
172. Institute for Community Medicine, University Medicine Greifswald, Greifswald, Germany.
173. Department of Gerontology and Geriatrics, Leiden University Medical Center, Leiden, the Netherlands.
174. Dasman Diabetes Institute, Dasman, Kuwait.
175. Chronic Disease Prevention Unit, National Institute for Health and Welfare, Helsinki, Finland.
176. Department of Public Health, University of Helsinki, Helsinki, Finland.
177. Saudi Diabetes Research Group, King Abdulaziz University, Jeddah, Saudi Arabia.
178. Department of Internal Medicine, Erasmus MC, Rotterdam, the Netherlands.
179. Research Institute for Primordial Prevention of Non-communicable Disease, Isfahan University of Medical Sciences, Isfahan, Iran.
180. Interfaculty Institute for Genetics and Functional Genomics, University Medicine Greifswald, Greifswald, Germany.
181. Department of Internal Medicine, University Hospital, CHUV, Lausanne, Switzerland.
182. Experimental Genetics Division, Sidra Medical and Research Center, Doha, Qatar.
183. Centre for Population Health Sciences, Usher Institute of Population Health Sciences and Informatics, University of Edinburgh, Scotland, UK
184. Department of Biology, Faculty of Medicine, University of Split, Croatia.
185. The National Institute for Health Research Blood and Transplant Research Unit in Donor Health and Genomics, University of Cambridge, UK.
186. Division of Cardiology, University Hospital, Basel, Switzerland.
187. Division of Cardiology, Department of Medicine, McMaster University, Hamilton, Canada.
188. Institute of Genetic and Biomedical Research, National Research Council (CNR), Monserrato, Cagliari, Italy.
189. Department of Biomedical Sciences, University of Sassari, Sassari, Italy.
190. Institute of Clinical Medicine, Internal Medicine, University of Eastern Finland and Kuopio University Hospital, Kuopio, Finland.
191. Laboratory of Cardiovascular Science, NIA/NIH, Baltimore, MD, USA.
192. Department of Public Health and Primary Care, Leiden University Medical Center, Leiden, the Netherlands.
193. Labormedizinisches Zentrum Dr. Risch, Schaan, Liechtenstein.
194. Private University of the Principality of Liechtenstin, Triesen, Liechtenstein.
195. University Insitute of Clinical Chemistry, Inselspital, Bern University Hospital, University of Bern, Bern, Switzerland.
196. Department of Cardiology, University of Groningen, University Medical Center Groningen, Groningen, The Netherlands.
197. Center for Genomic Medicine, Massachusetts General Hospital, Boston, MA, USA.
198. Cardiovascular Research Center, Massachusetts General Hospital, Boston, MA, USA.
201. Cardiovascular Health Research Unit, Departments of Medicine, Epidemiology and Health Services, University of Washington, Seattle, WA, USA.
202. Kaiser Permanente Washington Health Research Institute, Seattle, WA, USA.
203. National Institute for Health Research Imperial Biomedical Research Centre, Imperial College Healthcare NHS Trust and Imperial College London, London, UK.
204. UK Dementia Research Institute (UK DRI) at Imperial College London, London, UK
205. Health Data Research-UK London substantive site, London, U.K

## International Headache Genetic Consortium (IHGC)

Padhraig Gormley^1–4,81^, Verneri Anttila^2,3,5,81^, Bendik S Winsvold^6–8^, Priit Palta^9^, Tonu Esko^2,10,11^, Tune H Pers^2,11–13^, Kai-How Farh^2,5,14^, Ester Cuenca-Leon^1–3,15^, Mikko Muona^9,16–18^, Nicholas A Furlotte^19^, Tobias Kurth^20,21^, Andres Ingason^22^, George McMahon^23^, Lannie Ligthart^24^, Gisela M Terwindt^25^, Mikko Kallela^26^, Tobias M Freilinger^27,28^, Caroline Ran^29^, Scott G Gordon^30^, Anine H Stam^25^, Stacy Steinberg^22^, Guntram Borck^31^, Markku Koiranen^32^, Lydia Quaye^33^, Hieab H H Adams^34,35^, Terho Lehtimäki^36^, Antti-Pekka Sarin^9^, Juho Wedenoja^37^, David A Hinds^19^, Julie E Buring^21,38^, Markus Schürks^39^, Paul M Ridker^21,38^, Maria Gudlaug Hrafnsdottir^40^, Hreinn Stefansson^22^, Susan M Ring^23^, Jouke-Jan Hottenga^24^, Brenda W J H Penninx^41^, Markus Färkkilä^26^, Ville Artto^26^, Mari Kaunisto^9^, Salli Vepsäläinen^26^, Rainer Malik^28^, Andrew C Heath^42^, Pamela A F Madden^42^, Nicholas G Martin^30^, Grant W Montgomery^30^, Mitja I Kurki^1–3,9,43^, Mart Kals^10^, Reedik Mägi^10^, Kalle Pärn^10^, Eija Hämäläinen^9^, Hailiang Huang^2,3,5^, Andrea E Byrnes^2,3,5^, Lude Franke^44^, Jie Huang^4^, Evie Stergiakouli^23^, Phil H Lee^1–3^, Cynthia Sandor^45^, Caleb Webber^45^, Zameel Cader^46,47^, Bertram Muller-Myhsok^48,76,93^, Stefan Schreiber^49^, Thomas Meitinger^50,51^, Johan G Eriksson^52,53^, Veikko Salomaa^53^, Kauko Heikkilä^54^, Elizabeth Loehrer^34,55^, Andre G Uitterlinden^56^, Albert Hofman^34^, Cornelia M van Duijn^34^, Lynn Cherkas^33^, Linda M Pedersen^6^, Audun Stubhaug^57,58^, Christopher S Nielsen^57,59^, Minna Männikkö^32^, Evelin Mihailov^10^, Lili Milani^10^, Hartmut Göbel^60^, Ann-Louise Esserlind^61^, Anne Francke Christensen^61^, Thomas Folkmann Hansen^62^, Thomas Werge^63–65^, International Headache Genetics Consortium^66^, Jaakko Kaprio^9,37,67^, Arpo J Aromaa^53^, Olli Raitakari^68,69^, M Arfan Ikram^34,35,70^, Tim Spector^33^, Marjo-Riitta Järvelin^32,71–73^, Andres Metspalu^10^, Christian Kubisch^74^, David P Strachan^75^, Michel D Ferrari^25^, Andrea C Belin^29^, Martin Dichgans^28,76^, Maija Wessman^9,16^, Arn M J M van den Maagdenberg^25,77^, John-Anker Zwart^6–8^, Dorret I Boomsma^24^, George Davey Smith^23^, Kari Stefansson^22,78^, Nicholas Eriksson^19^, Mark J Daly^2,3,5^, Benjamin M Neale^2,3,5,82^, Jes Olesen^61,82^, Daniel I Chasman^21,38,82^, Dale R Nyholt^79,82^ & Aarno Palotie1–5,9,80,82

^1^Psychiatric and Neurodevelopmental Genetics Unit, Massachusetts General Hospital and Harvard Medical School, Boston, Massachusetts, USA. ^2^Medical and Population Genetics Program, Broad Institute of MIT and Harvard, Cambridge, Massachusetts, USA. ^3^Stanley Center for Psychiatric Research, Broad Institute of MIT and Harvard, Cambridge, Massachusetts, USA. ^4^Wellcome Trust Sanger Institute, Wellcome Trust Genome Campus, Hinxton, UK. ^5^Analytic and Translational Genetics Unit, Massachusetts General Hospital and Harvard Medical School, Boston, Massachusetts, USA. ^6^FORMI, Oslo University Hospital, Oslo, Norway. ^7^Department of Neurology, Oslo University Hospital, Oslo, Norway. ^8^Institute of Clinical Medicine, University of Oslo, Oslo, Norway. ^9^Institute for Molecular Medicine Finland (FIMM), University of Helsinki, Helsinki, Finland. ^10^Estonian Genome Center, University of Tartu, Tartu, Estonia. ^11^Division of Endocrinology, Boston Children’s Hospital, Boston, Massachusetts, USA. ^12^Department of Epidemiology Research, Statens Serum Institut, Copenhagen, Denmark. ^13^Novo Nordisk Foundation Center for Basic Metabolic Research, University of Copenhagen, Copenhagen, Denmark. ^14^Illumina, San Diego, California, USA. ^15^Pediatric Neurology, Vall d’Hebron Research Institute, Barcelona, Spain. ^16^Folkhälsan Institute of Genetics, Helsinki, Finland. ^17^Neuroscience Center, University of Helsinki, Helsinki, Finland. ^18^Molecular Neurology Research Program, Research Programs Unit, University of Helsinki, Helsinki, Finland. ^19^23andMe, Inc., Mountain View, California, USA. ^20^Institute of Public Health, Charité– Universitätsmedizin Berlin, Berlin, Germany. ^21^Division of Preventive Medicine, Brigham and Women’s Hospital, Boston, Massachusetts, USA. ^22^deCODE Genetics, Reykjavik, Iceland. ^23^Medical Research Council (MRC) Integrative Epidemiology Unit, University of Bristol, Bristol, UK. ^24^Department of Biological Psychology, Vrije Universiteit, Amsterdam, the Netherlands. ^25^Department of Neurology, Leiden University Medical Centre, Leiden, the Netherlands. ^26^Department of Neurology, Helsinki University Central Hospital, Helsinki, Finland. ^27^Department of Neurology and Epileptology, Hertie-Institute for Clinical Brain Research, University of Tuebingen, Tuebingen, Germany. ^28^Institute for Stroke and Dementia Research, Klinikum der Universität München, Ludwig-Maximilians-Universität München, Munich, Germany. ^29^Department of Neuroscience, Karolinska Institutet, Stockholm, Sweden. ^30^Department of Genetics and Computational Biology, QIMR Berghofer Medical Research Institute, Brisbane, Queensland, Australia. ^31^Institute of Human Genetics, Ulm University, Ulm, Germany. ^32^Center for Life Course Epidemiology and Systems Medicine, University of Oulu, Oulu, Finland. ^33^Department of Twin Research and Genetic Epidemiology, King’s College London, London, UK. ^34^Department of Epidemiology, Erasmus University Medical Center, Rotterdam, the Netherlands. ^35^Department of Radiology, Erasmus University Medical Center, Rotterdam, the Netherlands. ^36^Department of Clinical Chemistry, Fimlab Laboratories, School of Medicine, University of Tampere, Tampere, Finland. ^37^Department of Public Health, University of Helsinki, Helsinki, Finland. ^38^Harvard Medical School, Boston, Massachusetts, USA. ^39^Department of Neurology, University Duisburg–Essen, Essen, Germany. ^40^Landspitali University Hospital, Reykjavik, Iceland. ^41^Department of Psychiatry, VU University Medical Centre, Amsterdam, the Netherlands. ^42^Department of Psychiatry, Washington University School of Medicine, St. Louis, Missouri, USA. ^43^Department of Neurosurgery, NeuroCenter, Kuopio University Hospital, Kuopio, Finland. ^44^Department of Genetics, University Medical Center Groningen, University of Groningen, Groningen, the Netherlands. ^45^MRC Functional Genomics Unit, Department of Physiology, Anatomy & Genetics, Oxford University, Oxford, UK. ^46^Nuffield Department of Clinical Neuroscience, University of Oxford, Oxford, UK. ^47^Oxford Headache Centre, John Radcliffe Hospital, Oxford, UK. ^48^Max Planck Institute of Psychiatry, Munich, Germany. ^49^Institute of Clinical Molecular Biology, Christian Albrechts University, Kiel, Germany. ^50^Institute of Human Genetics, Helmholtz Zentrum München, Neuherberg, Germany. ^51^Institute of Human Genetics, Technische Universität München, Munich, Germany. ^52^Department of General Practice and Primary Health Care, University of Helsinki and Helsinki University Hospital, Helsinki, Finland. ^53^National Institute for Health and Welfare, Helsinki, Finland. ^54^Institute of Clinical Medicine, University of Helsinki, Helsinki, Finland. ^55^Department of Environmental Health, Harvard T.H. Chan School of Public Health, Boston, Massachusetts, USA. ^56^Department of Internal Medicine, Erasmus University Medical Center, Rotterdam, the Netherlands. ^57^Department of Pain Management and Research, Oslo University Hospital, Oslo, Norway. ^58^Medical Faculty, University of Oslo, Oslo, Norway. ^59^Department of Ageing and Health, Norwegian Institute of Public Health, Oslo, Norway. ^60^Kiel Pain and Headache Center, Kiel, Germany. ^61^Danish Headache Center, Department of Neurology, Rigshospitalet, Glostrup Hospital, University of Copenhagen, Copenhagen, Denmark. ^62^Institute of Biological Psychiatry, Mental Health Center Sct. Hans, University of Copenhagen, Roskilde, Denmark. ^63^Institute of Biological Psychiatry, MHC Sct. Hans, Mental Health Services Copenhagen, Copenhagen, Denmark. ^64^Institute of Clinical Sciences, Faculty of Medicine and Health Sciences, University of Copenhagen, Copenhagen, Denmark. ^65^iPSYCH—The Lundbeck Foundation Initiative for Integrative Psychiatric Research, Copenhagen, Denmark. ^66^http://www.headachegenetics.org/. ^67^Department of Health, National Institute for Health and Welfare, Helsinki, Finland. ^68^Research Centre of Applied and Preventive Cardiovascular Medicine, University of Turku, Turku, Finland. ^69^Department of Clinical Physiology and Nuclear Medicine, Turku University Hospital, Turku, Finland. ^70^Department of Neurology, Erasmus University Medical Center, Rotterdam, the Netherlands. ^71^Department of Epidemiology and Biostatistics, MRC Health Protection Agency (HPE) Centre for Environment and Health, School of Public Health, Imperial College London, London, UK. ^72^Biocenter Oulu, University of Oulu, Oulu, Finland. ^73^Unit of Primary Care, Oulu University Hospital, Oulu, Finland. ^74^Institute of Human Genetics, University Medical Center Hamburg-Eppendorf, Hamburg, Germany. ^75^Population Health Research Institute, St George’s, University of London, London, UK. ^76^Munich Cluster for Systems Neurology (SyNergy), Munich, Germany. ^77^Department of Human Genetics, Leiden University Medical Centre, Leiden, the Netherlands. ^78^Faculty of Medicine, University of Iceland, Reykjavik, Iceland. ^79^Statistical and Genomic Epidemiology Laboratory, Institute of Health and Biomedical Innovation, Queensland University of Technology, Kelvin Grove, Queensland, Australia. ^80^Department of Neurology, Massachusetts General Hospital, Boston, Massachusetts, USA. ^81^These authors contributed equally to this work. ^82^These authors jointly supervised this work.

## International Stroke Genetics Consortium (ISGC) Intracranial Aneurysm Working Group

Mark K. Bakker^1^, Romain Bourcier^2,3^, Robin G. Walters^4,5^, Rainer Malik^6^, Martin Dichgans^6,7,8^, Muralidharan Sargurupremraj^9,10^, Turgut Tatlisumak^11^, Stéphanie Debette^9,10^, Gabriel J.E. Rinkel^1^, Bradford B. Worrall^12^, Joanna Pera^13^, Agnieszka Slowik^13^, Joseph P. Broderick^14^, David J. Werring^15^, Daniel Woo^14^, Philippe Bijlenga^16^, Yoichiro Kamatani^17^, Ynte M. Ruigrok^1^

^1^Department of Neurology and Neurosurgery, University Medical Center Utrecht Brain Center, Utrecht University, Utrecht, The Netherlands. ^2^Université de Nantes, CHU Nantes, INSERM, CNRS, l’institut du thorax, Nantes, France. ^3^CHU Nantes, Department of Neuroradiology, Nantes, France. ^4^Clinical Trial Service Unit and Epidemiological Studies Unit, Nuffield Department of Population Health, University of Oxford, Oxford, U.K. ^5^Medical Research Council Population Health Research Unit, University of Oxford, Oxford, U.K. ^6^Institute for Stroke and Dementia Research, University Hospital, Ludwig-Maximilians-University, Munich. ^7^Munich Cluster for Systems Neurology (SyNergy), Munich, Germany. ^8^Deutsches Zentrum für Neurodegenerative Erkrankungen (DZNE), Munich, Germany. ^9^INSERM U1219 Bordeaux Population Health Research Center, University of Bordeaux, Bordeaux, France. ^10^Department of Neurology, Institute for Neurodegenerative Disease, Bordeaux University Hospital, Bordeaux, France. ^11^Department of Clinical Neuroscience at Institute of Neuroscience and Physiology, University of Gothenburg, Sweden. ^12^Departments of Neurology and Public Health Sciences, University of Virginia School of Medicine, Charlottesville, VA, USA. ^13^Department of Neurology, Faculty of Medicine, Jagiellonian University Medical College, ul. Botaniczna 3, 31-503, Krakow, Poland. ^14^University of Cincinnati College of Medicine, Cincinnati, OH, USA. ^15^Stroke Research Centre, University College London Queen Square Institute of Neurology, London, UK. ^16^Neurosurgery Division, Department of Clinical Neurosciences, Faculty of Medicine, Geneva University Hospitals, Geneva, Switzerland. ^17^Graduate School of Frontier Sciences, The University of Tokyo, Tokyo, Japan.

