## Supplementary Data for "Genetic risk score for intracranial aneurysms to predict aneurysmal subarachnoid hemorrhage and identify associations with patient characteristics"

### Supplementary Methods

#### Association data preprocessing

For each set of summary statistics obtained from sources described in Supplementary Table 1, the following pre-processing steps were taken. Missing alternate alleles, minor allele frequencies (MAF) and chromosomal positions were annotated from the haplotype reference consortium (HRC) release 1.1 on reference genome GRCh37. If single-nucleotide polymorphism (SNP) beta, standard error (SE), and/or P-value were not available, these were calculated from the available or annotated effect size, SE, P-value, Z-score, sample size, and MAF. Effective sample size and per-SNP sample sizes were used where available.

#### Cohort descriptions

**UK Biobank.**[1] This dataset was used to determine the optimal trait-level GRSs, and to combine trait-level GRSs into a metaGRS. Persons with an unruptured intracranial aneurysm (UIA) or aneurysmal subarachnoid hemorrhage (ASAH), both assessed by ICD-10 codes, were selected as intracranial aneurysm (IA) cases. Persons with a non-aneurysmal subarachnoid hemorrhage, with another ancestral background than white British, or with a diagnosis of autosomal dominant polycystic kidney disease, Ehlers-Danlos disease or Marfan syndrome, were excluded. SNPs with MAF < 0.005 or imputation INFO-score < 0.60 were excluded. This resulted in 1,161 intracranial aneurysm (IA) patients, of which 959 had an ASAH, 202 with UIA, and 408,553 controls.

**HUNT study.** This dataset was used to assess predictive performance of the metaGRS for ASAH hazard and IA presence. Detailed information about inclusion criteria, diagnosis and genotyping has been described before.[2, 3] In brief, UIA and ASAH diagnosis was done by ICD-10 codes I67.1 and I60. A total of 828 IA patients, of which 318 with ASAH, and 68,568 without any IA were included. For the analyses studying ASAH, the UIA cases were included as controls, resulting in 69,078 controls. Genotyping was done using Illumina HumanCoreExome platforms.

**Phenotype cohort.** From a subset of the largest GWAS on IA, detailed phenotype information was obtained.[4] The phenotype cohort was used to assess which phenotypes were associated with genetic predisposition for IA. Persons without genetic data were excluded. Phenotypic information was obtained in 5,560 IA patients of which 3,916 with ASAH from 18 European cohorts. In 1,544 patients multiple IAs were found. Phenotypes were obtained from imaging data or surgical exploration.

#### Genetic correlation

To select traits to include in elastic net regression, we calculated genetic correlation between IA and the respective trait using linkage disequilibrium score regression (LDSC).[5] HapMap v3 linkage disequilibrium (LD) scores were used as reference panel. Only SNPs with minor allele frequency > 5% were included, and an LD window of 200 kb was selected. Traits with a genetic correlation P-value < 0.05 were selected to create a trait-level GRS.

#### Training the trait-level genetic risk scores

Eleven trait-level GRSs were calculated for each trait according to three methods: LD-based clumping with 9 LD thresholds, summary statistics-based best linear unbiased predictor (s-BLUP)[6] and summary statistics-based bayes R (s-Bayes R)[7]. No P-value threshold was used to select SNPs for any of the models. Control samples included in the largest IA GWAS[4] (stratum sNL2) were used as LD reference panel. Clumping was done to exclude LD-correlated SNPs, while retaining the SNPs with lowest P-values. This was done using plink v1.9.[8] LD r-squared thresholds ranging from 0.1 to 0.9 (interval 0.1) were used. s-BLUP scales SNP weights to provide LD-adjusted linear predictors. s-BLUP lambda was calculated by dividing the number of SNPs in common between summary statistics and LD reference panel by the following: the inverse of the SNP-based heritability estimate as assessed by LDSC, minus 1. For heritability estimation, the same SNPs and settings were used as for genetic correlation estimation. s-Bayes R uses a Bayesian approach to calculate LD-adjusted SNP effects suitable for genetic risk prediction. For s-Bayes R, a sparse LD matrix was created with SNPs in common between the LD reference panel and summary statistics. An interpolated genetic map with centimorgan positions was used (ftp://ftp.1000genomes.ebi.ac.uk/vol1/ftp/technical/working/20130507_omni_recombination_rates/). SNP weights were then calculated with a pi vector of 0.95, 0.02, 0.02 and 0.01 and a gamma vector of 0, 1, 10 and

100, while allowing for unscaled genotypes. A burn-in of 10,000 iterations and total chain of 20,000 iterations were selected.

For each of the 11 models (9 clumping thresholds, s-BLUP, and s-Bayes R), model performance was assessed in the UK Biobank cohort by logistic regression in R (function *glm*), using sex (in model analyzing whole cohort) and 10 genetic principal components as covariates. The GRS with the highest Nagelkerke pseudo R-squared was selected for elastic net regression, leaving each trait with three optimal trait-GRS models: one for the whole cohort, one for men only and one for women only.

**Association of trait-level GRSs with their respective traits**

We assessed whether trait-level GRSs were associated with their respective traits in the UK Biobank. Trait-level SNP weights as included in the metaGRS were used. Traits included in the metaGRS, for which a representative phenotype was available in the UK Biobank, with at least 100 cases (for binary traits) were selected for this analysis (Supplementary Table 6). ICD-10 codes for hospitalization, or death record were used for binary traits. Individual-level trait-level GRSs were calculated using *plink (v2.0)* function *--score*, only including SNPs with INFO-score above 0.8. Each trait-level GRS was transformed to zero mean, unit variance. For binary traits, a logistic regression with trait-level GRS as independent variable, respective trait as dependent variable, and sex and age as covariates was performed in R. A model excluding the trait-level GRS was used to assess the improvement of the area under the receiver operator characteristic curve upon adding the trait-level GRS. For quantitative traits a linear regression was performed using the same variables. Added value of the trait-level GRS was assessed by comparing r-squared value between the models including and excluding the trait-level GRS. For age at menarche, only women were included, and sex was therefore not included as covariate.

**Creating the metaGRS from trait-level GRSs**

Individual level trait-level GRSs (of the optimal trait-level GRSs) were calculated using *plink (v1.9)* function *--score* in the UK Biobank cohort, only including SNPs with INFO-score above 0.8. All trait-level GRSs were then standardized to mean 0 and standard deviation 1. Elastic net regression was performed on all trait-level GRSs combined using R function *glmnet* using covariates sex (in model analyzing whole cohort) and 10 genetic principal components. Ten-fold cross-validation was performed to optimize the area under the curve and thereby obtain the trait-level GRS elastic net weights. For each trait, per-SNP weights from the trait-level GRS models were multiplied by the elastic net weight for that trait and divided by the population GRS standard deviation for that trait in the UK Biobank cohort prior to standardizing. These per-SNP weights were summed over traits, creating per-SNP weights of the metaGRS. The IA trait-level SNP weights after elastic net regression were also used as a IA-only GRS to compare metaGRS and IA-only GRS performance.

**Covariate selection for metaGRS prediction assessment**

Variables to include in the prediction assessment model were selected by logistic regression (function *glm* in R) with IA as outcome in the UK Biobank. Variables with a statistically significant effect in the UK Biobank (P-value < 0.05), while correcting for sex and age (except when testing these), were included in the model. Variables tested were: sex, age, systolic blood pressure, diastolic blood pressure, alcoholic drinks per week, average smoking packs per day since age 16. Alcoholic drinks per week was not statistically significant as predictor, and diastolic blood pressure was excluded because of the strong correlation with, but less statistically significant effect than, systolic blood pressure. For each variable, we determined whether to use it as linear predictor, cubic spline, or polynomial spline, by optimizing the Akaike information criterion in a logistic regression model with IA as outcome, the respective variable as predictor, and covariates age and sex. We varied splines from 3-10 knots. Age, smoking pack years proportional to lifespan, and SBP were used as polynomial spline with 3 knots, while metaGRS was used as a linear predictor.

**Creating a leave-one-out metaGRS for association with patient characteristics**

To account for sample overlap between IA GWAS and the phenotype cohort, nine sets of IA GWAS summary statistics were created excluding each stratum in a leave-one-out manner. Stratum sUK, including the UK Biobank, was left out in every set due to overlap with the metaGRS training set (as this stratum was also left out when creating the main metaGRS used for prediction of ASAH hazard and IA presence). Nine metaGRS versions were created by elastic net regression using each leave-one-out IA GWAS once, selecting the same optimal trait-level GRSs (LD clumping, s-Bayes R, or s-BLUP) as used for the main metaGRS. Individual-level

GRS of all patients were calculated with plink option --*score* using the per-SNP weights of the metaGRS leaving out the patients' respective stratum, as SNP scores.

**Calculating predicted cumulative incidence**

Cox regression was performed using R function *coxph*. Baseline hazard at age 75 was calculated using R function *basehaz*. Predicted cumulative incidence of a person of age 75 with varying sex and metaGRS was calculated by multiplying the baseline hazard with natural exponentiation of the log-hazard ratio obtained by the *predict* function.

**Association between metaGRS and patient characteristics**

We calculated the effect of various phenotypes (hypertension, smoking status, IA multiplicity, rupture status, age at ASAH, aneurysmal size at rupture, family history, and IA locations) on the metaGRS. To determine the effect of age at ASAH and aneurysmal size at rupture, only ASAH patients were included. Persons with an aneurysmal size below 1.5 times the interquartile range lower than the first quartile, or above 1.5 times the interquartile range higher than the third quartile were considered outliers and excluded from the aneurysmal size analysis. When assessing the effect of IA location, only patients with a single IA were included. For each phenotype, a generalized linear model was fitted with metaGRS as dependent variable, the phenotype of interest as independent variable, and cohort and sex as covariates. To obtain an interpretable effect size of metaGRS on age at ASAH, an additional analysis was done with age at ASAH as outcome and metaGRS as independent variable. Effect size of IA phenotypes on the metaGRS were transformed to unit variance by dividing the effect size by the standard deviation of the metaGRS among included samples. Odds ratios were then calculated for binary phenotypes by natural exponentiation.

To test whether the effect of a phenotype on the metaGRS remained when including other phenotypes, we used a stepwise selection model (R function *stepAIC* ). One model was used for each subset (all cases, single IA cases, and ASAH cases). Each model included sex, hypertension, smoking status, and cohort. Depending on the subset of cases, the model could also include IA multiplicity, rupture status, family history, and IA location at the ICA.

### Supplementary Figures

**Supplementary Figure 1. Predictive performance for aneurysmal subarachnoid hemorrhage of the metaGRS in women and men, alone.** A) metaGRS trained using an elastic net regression in women included the UK Biobank, and validated in women included in the HUNT study. B) The same as panel A, but trained and validated in men.

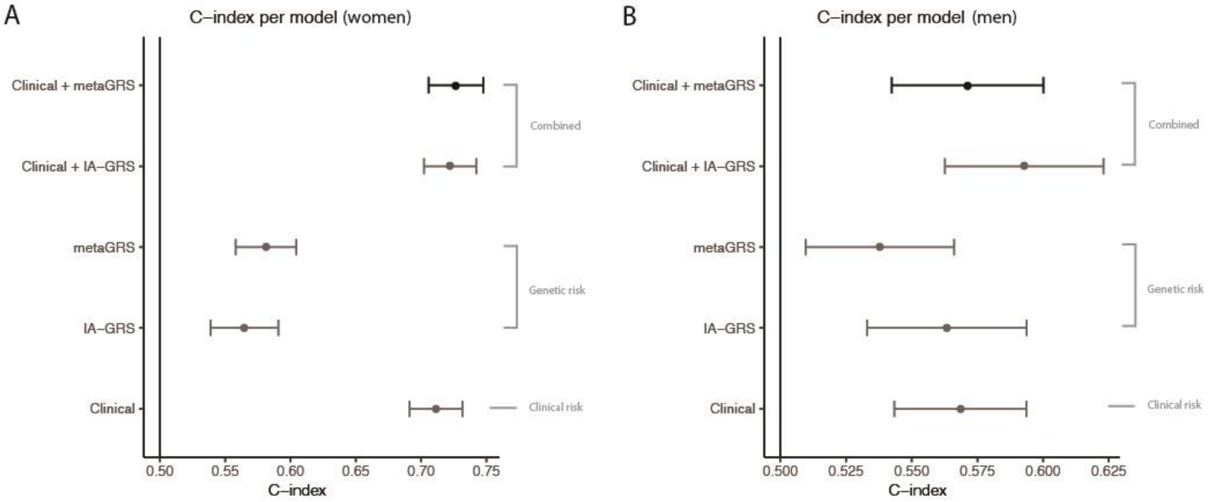

**Supplementary Figure 2. Predictive performance for intracranial aneurysm presence of the metaGRS.** metaGRS trained using an elastic net regression in men and women combined in the UK Biobank, and validated in the HUNT study. Reference: prediction model including age and sex only. Clinical: model including age + sex + systolic blood pressure (SBP) + smoking packs per day. metaGRS: model including age + sex + metaGRS. The models above the dashed line each leave out one variable from the full model which includes age + sex + SBP + smoking packs per day + metaGRS. Error bars denote 95% confidence interval ($CI_{95}$). AUC: area under the receiver operator characteristic curve.

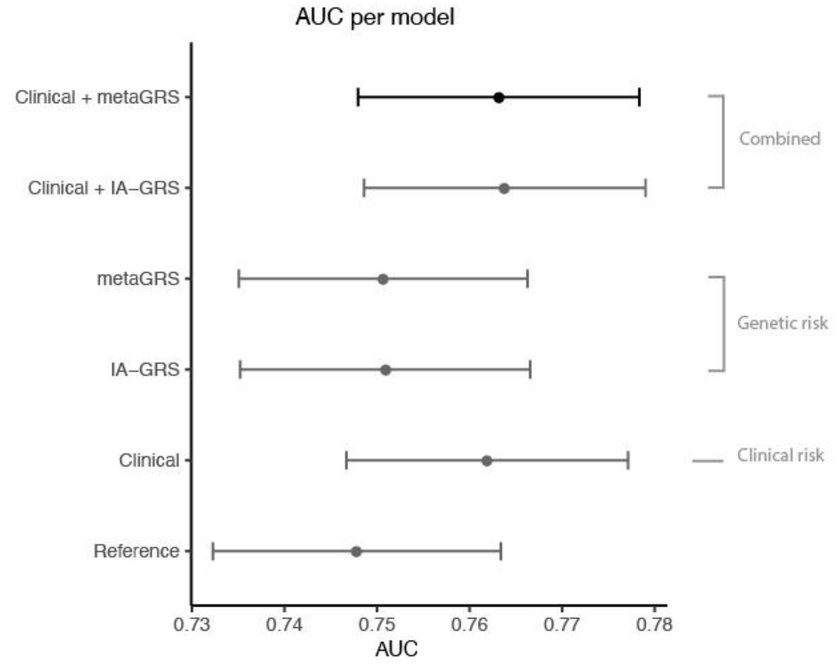

**Supplementary Figure 3. Association between metaGRS and intracranial aneurysm (IA, either ruptured or unruptured) location at the internal carotid artery (ICA) versus other locations.** A) Violin plot of the distribution of metaGRS in the phenotype cohort among persons with an IA at the ICA and the remaining group. Horizontal lines denote mean and mean ± 1 standard deviation. B) Box plots showing the distribution in each sub-cohort within the phenotype cohort. Boxes contain 25[th] to 75[th] percentile and denote the median with a horizontal line. Whiskers denote smallest value greater than 1.5 times the interquartile range below the 25[th] percentile, and largest value smaller than 1.5 times the interquartile range above the 75[th] percentile.

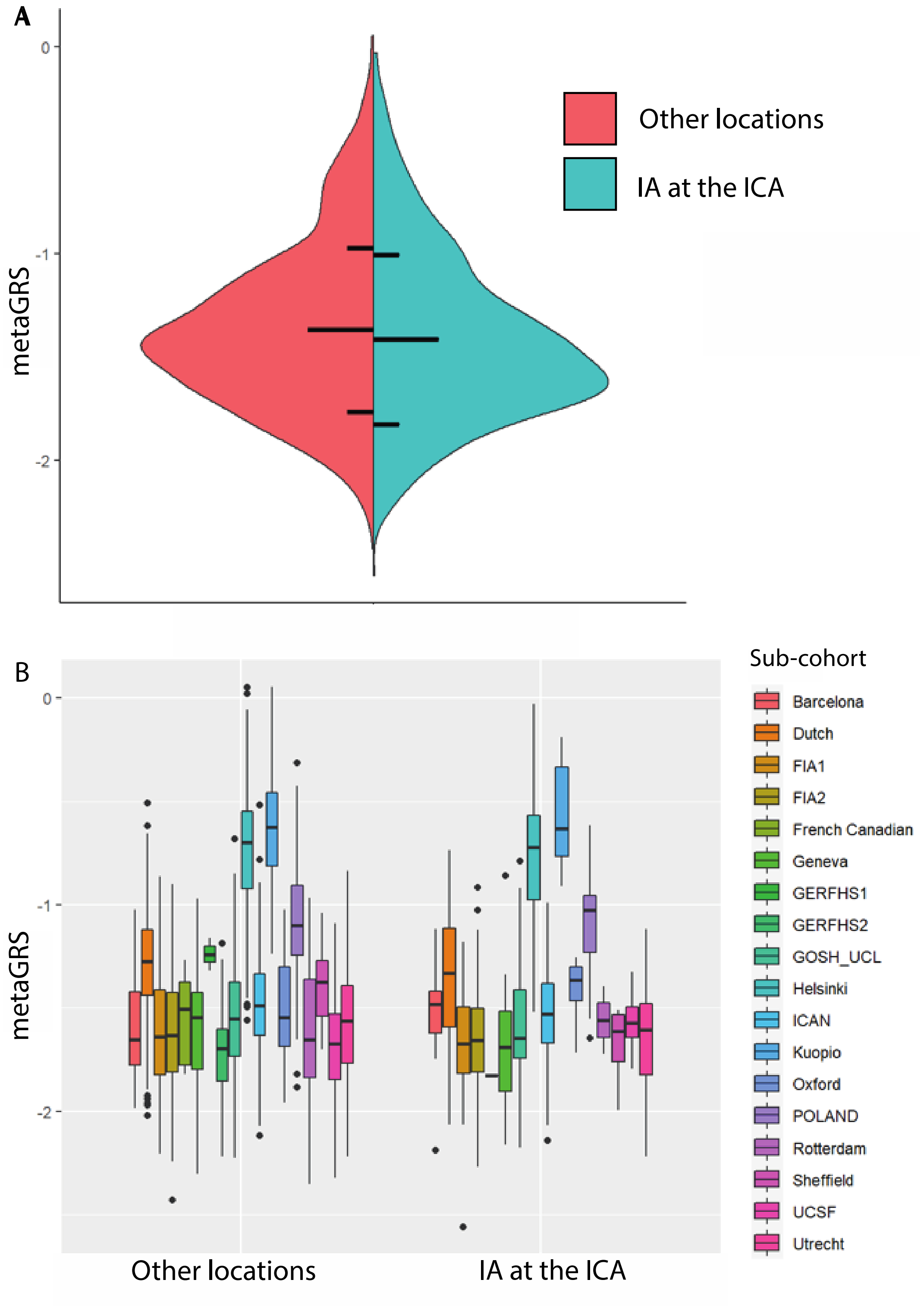

**Supplementary Figure 4. Association between metaGRS and intracranial aneurysm (IA, either ruptured or unruptured) location at the posterior communicating artery (PCOM) versus other locations.** A) Violin plot of the distribution of metaGRS in the phenotype cohort among persons with an IA at the PCOM and the remaining group. Horizontal lines denote mean and mean ± 1 standard deviation. B) Box plots showing the distribution in each sub-cohort within the phenotype cohort. Boxes contain 25th to 75th percentile and denote the median with a horizontal line. Whiskers denote smallest value greater than 1.5 times the interquartile range below the 25th percentile, and largest value smaller than 1.5 times the interquartile range above the 75th percentile.

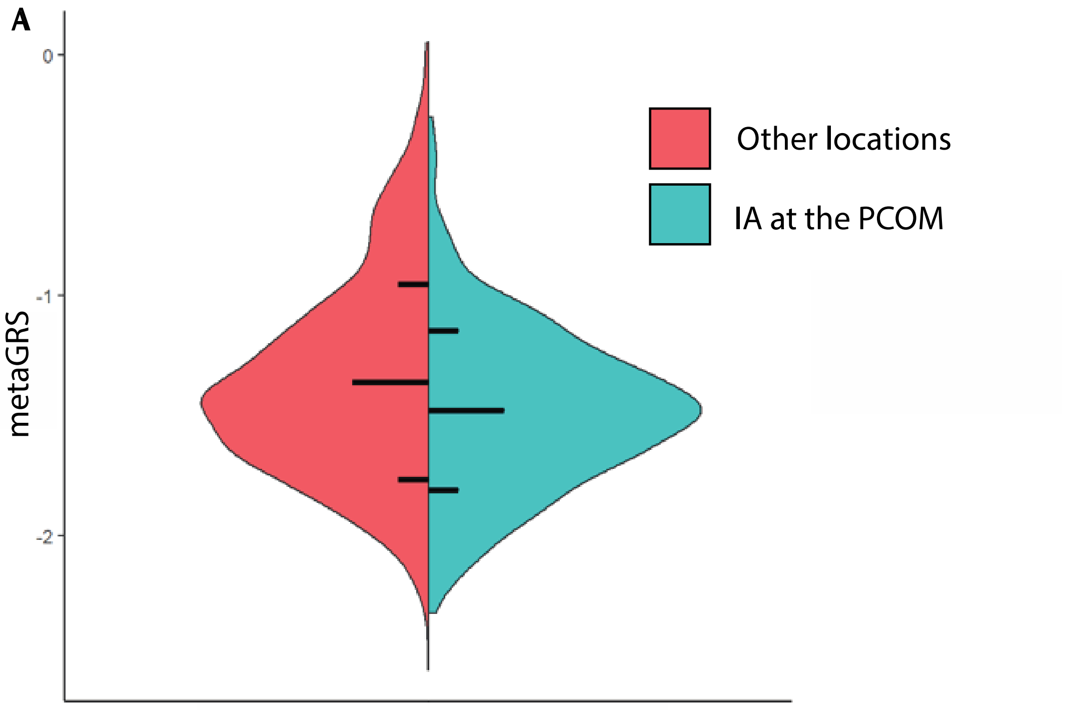

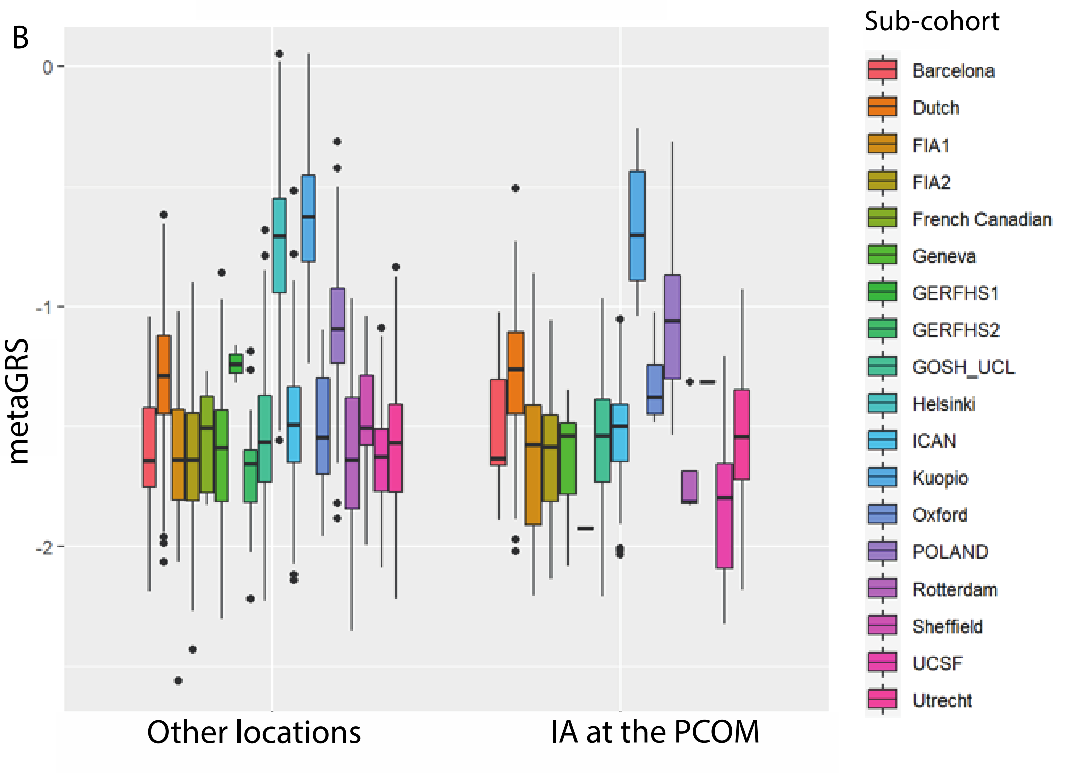

**Supplementary Figure 5. Association between metaGRS and intracranial aneurysm (IA, either ruptured or unruptured) location at the anterior cerebral arteries (ACA) versus other locations.** A) Violin plot of the distribution of metaGRS in the phenotype cohort among persons with an IA at the ACA and the remaining group. Horizontal lines denote mean and mean ± 1 standard deviation. B) Box plots showing the distribution in each sub-cohort within the phenotype cohort. Boxes contain 25th to 75th percentile and denote the median with a horizontal line. Whiskers denote smallest value greater than 1.5 times the interquartile range below the 25th percentile, and largest value smaller than 1.5 times the interquartile range above the 75th percentile.

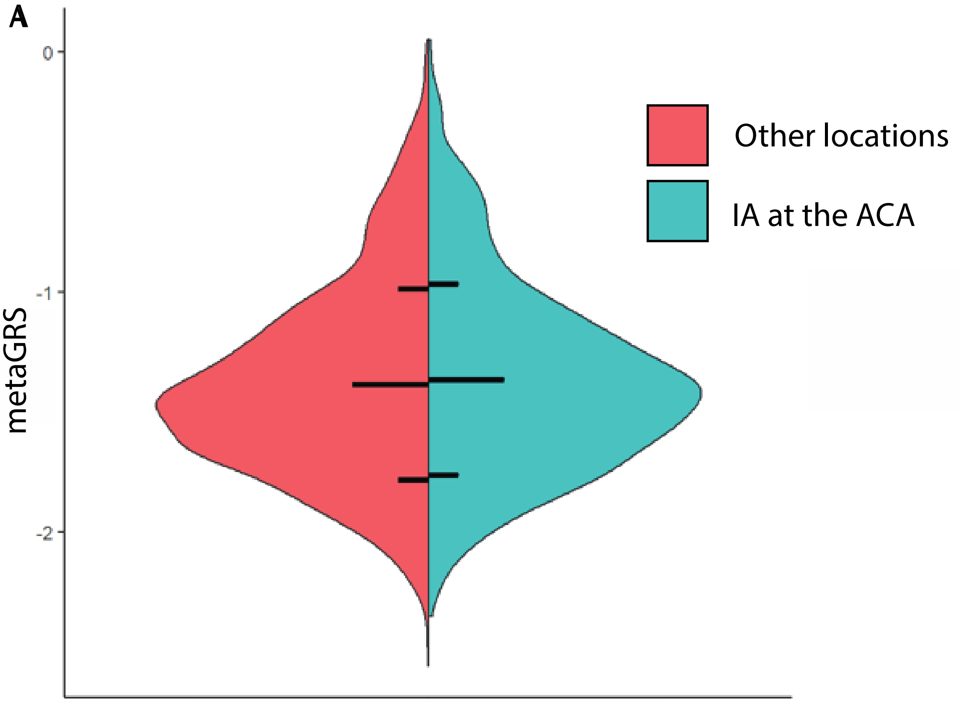

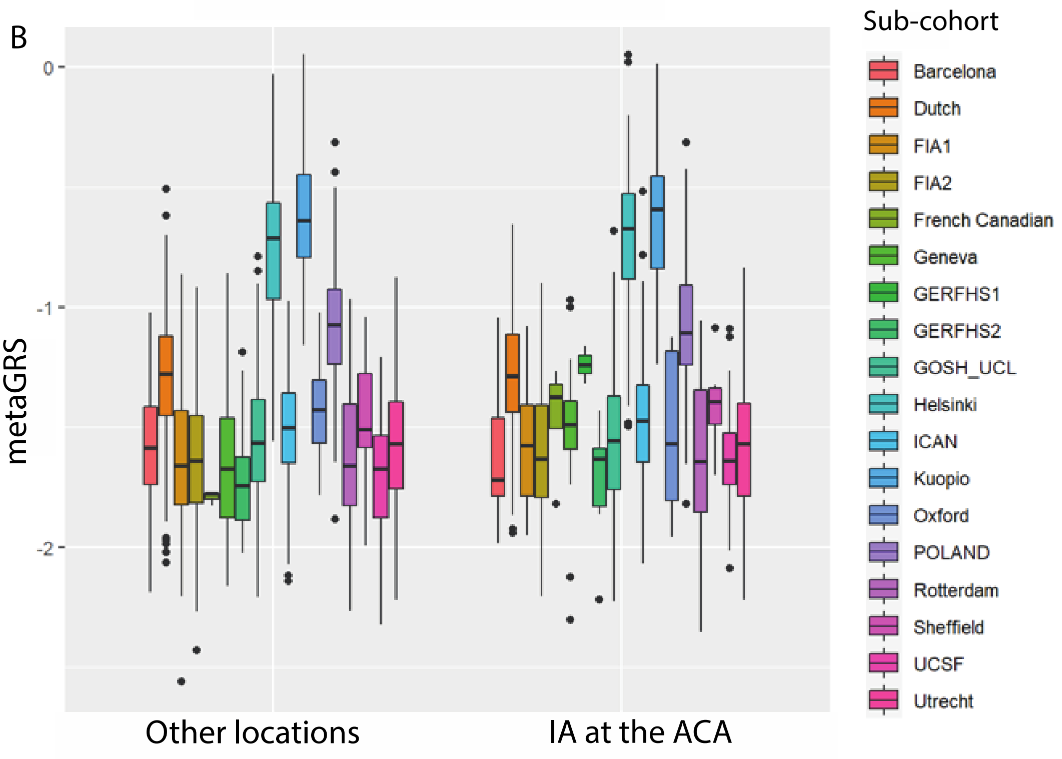

**Supplementary Figure 6. Association between metaGRS and intracranial aneurysm (IA, either ruptured or unruptured) location at the middle cerebral artery (MCA) versus other locations.** A) Violin plot of the distribution of metaGRS in the phenotype cohort among persons with an IA at the MCA and the remaining group. Horizontal lines denote mean and mean ± 1 standard deviation. B) Box plots showing the distribution in each sub-cohort within the phenotype cohort. Boxes contain 25th to 75th percentile and denote the median with a horizontal line. Whiskers denote smallest value greater than 1.5 times the interquartile range below the 25th percentile, and largest value smaller than 1.5 times the interquartile range above the 75th percentile.

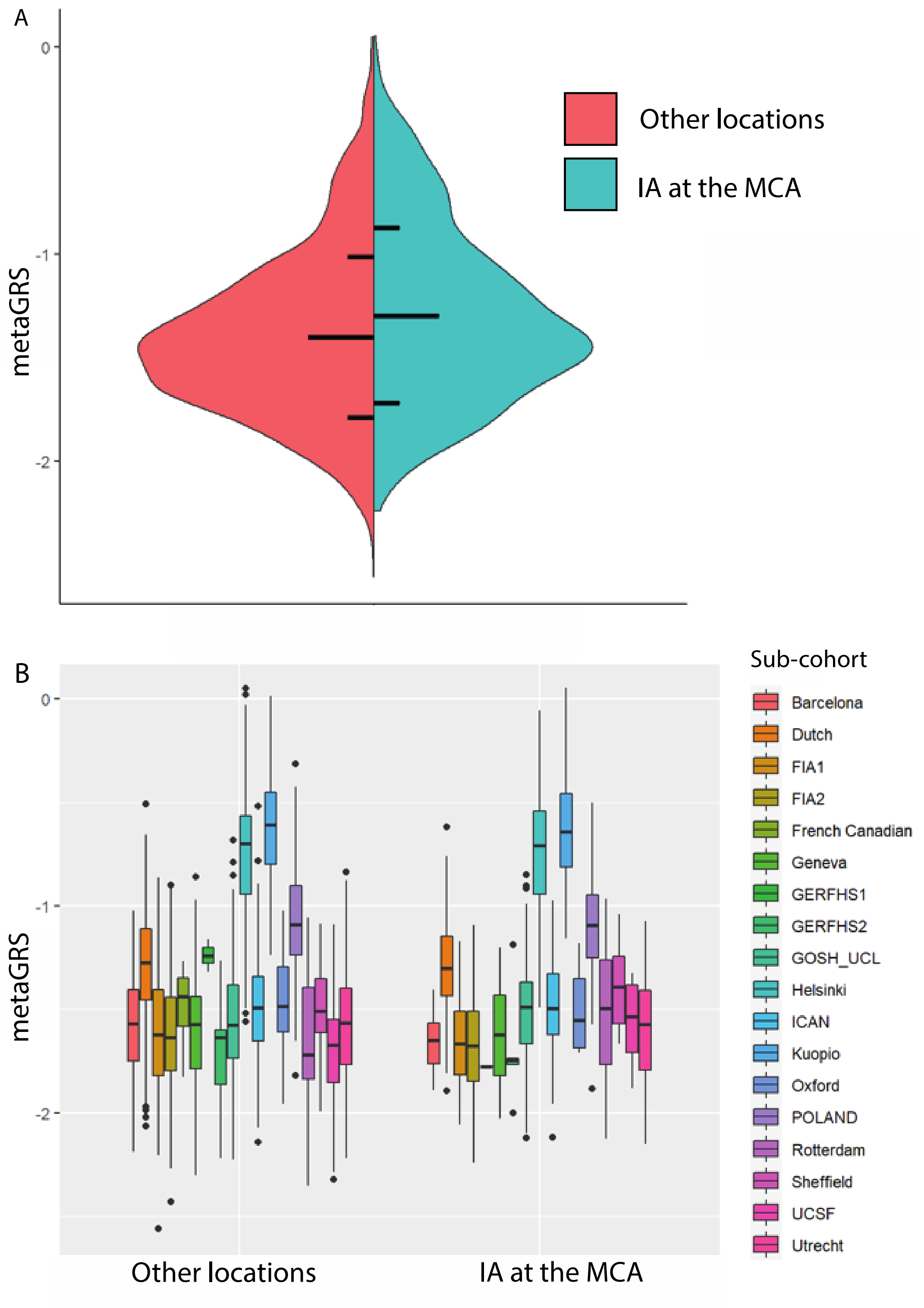

**Supplementary Figure 7. Association between metaGRS and intracranial aneurysm (IA, either ruptured or unruptured) location at the posterior circulation arteries (PC) versus other locations.** A) Violin plot of the distribution of metaGRS in the phenotype cohort among persons with an IA at the PC and the remaining group. Horizontal lines denote mean and mean ± 1 standard deviation. B) Box plots showing the distribution in each sub-cohort within the phenotype cohort. Boxes contain 25th to 75th percentile and denote the median with a horizontal line. Whiskers denote smallest value greater than 1.5 times the interquartile range below the 25th percentile, and largest value smaller than 1.5 times the interquartile range above the 75th percentile.

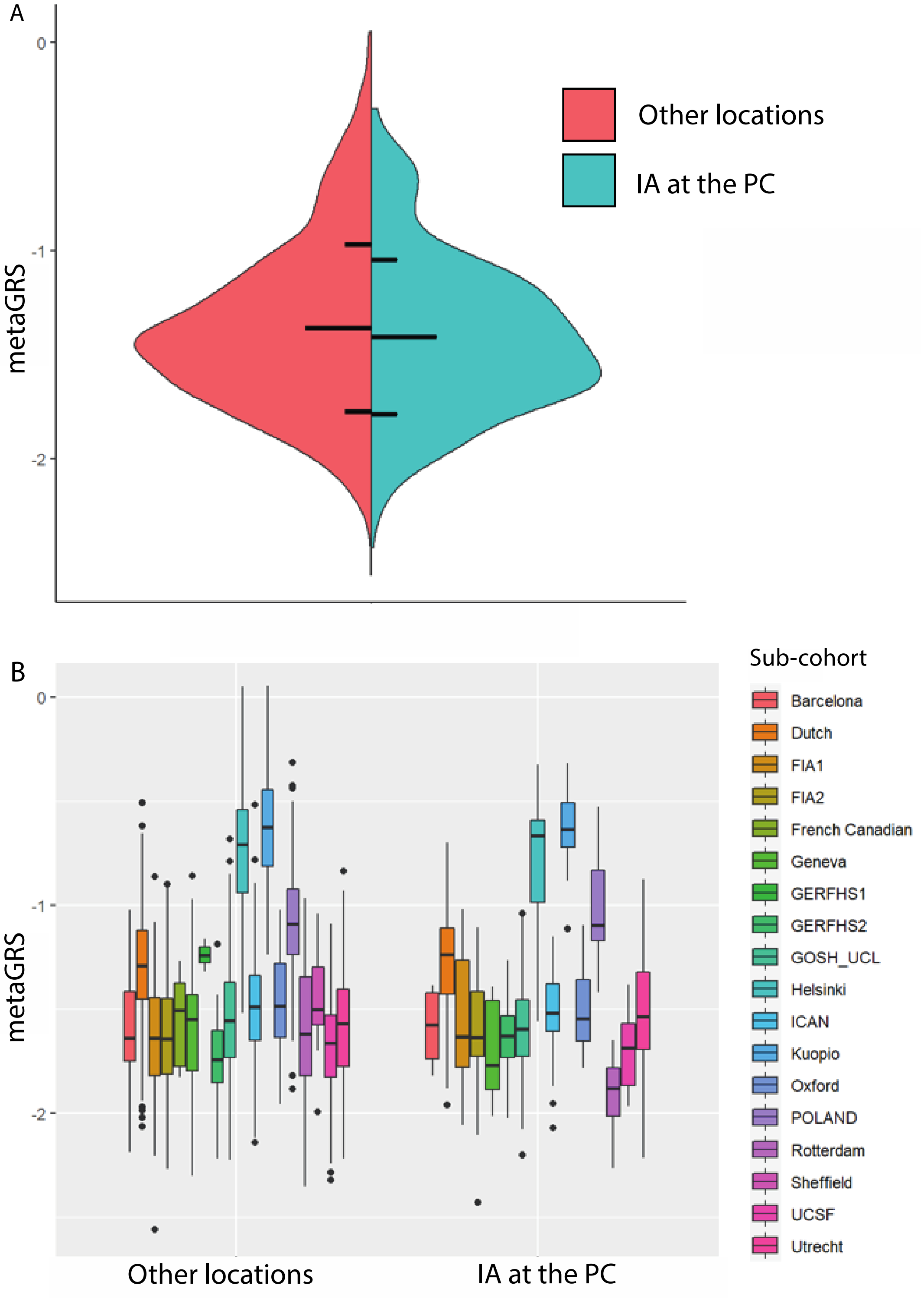

**Supplementary Figure 8. Association between metaGRS and aneurysmal subarachnoid hemorrhage (ASAH) from an intracranial aneurysm (IA) at the internal carotid artery (ICA) versus other locations.** A) Violin plot of the distribution of metaGRS in the phenotype cohort among persons with an ASAH at the ICA and the remaining group. Horizontal lines denote mean and mean ± 1 standard deviation. B) Box plots showing the distribution in each sub-cohort within the phenotype cohort. Boxes contain 25th to 75th percentile and denote the median with a horizontal line. Whiskers denote smallest value greater than 1.5 times the interquartile range below the 25th percentile, and largest value smaller than 1.5 times the interquartile range above the 75th percentile.

A

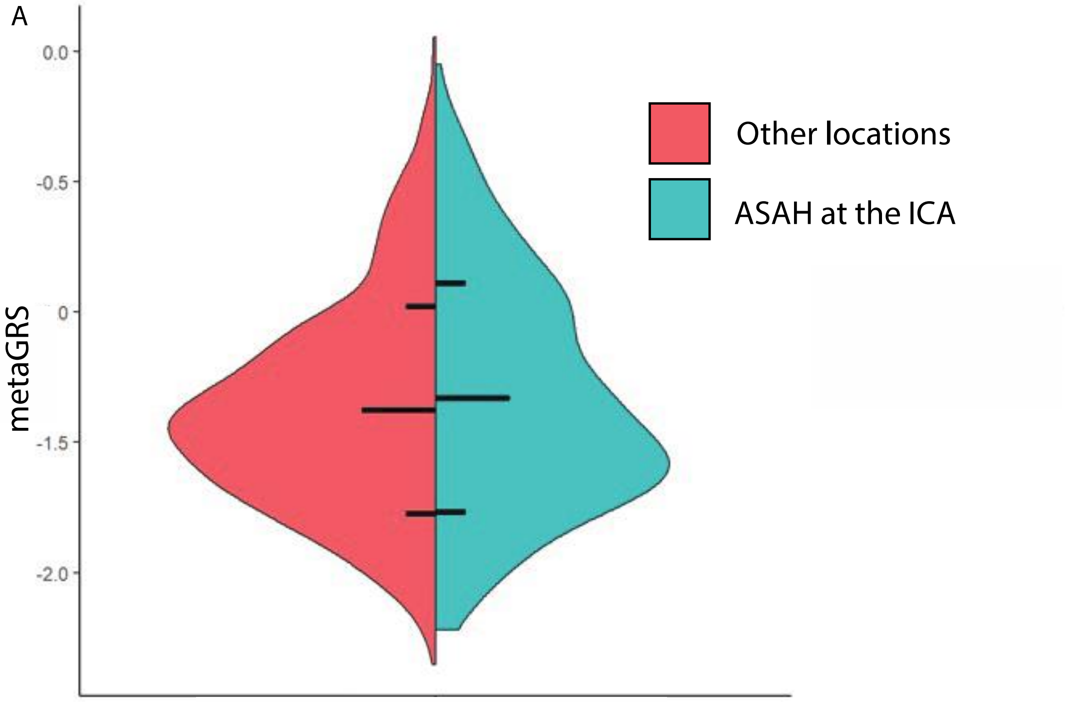

B

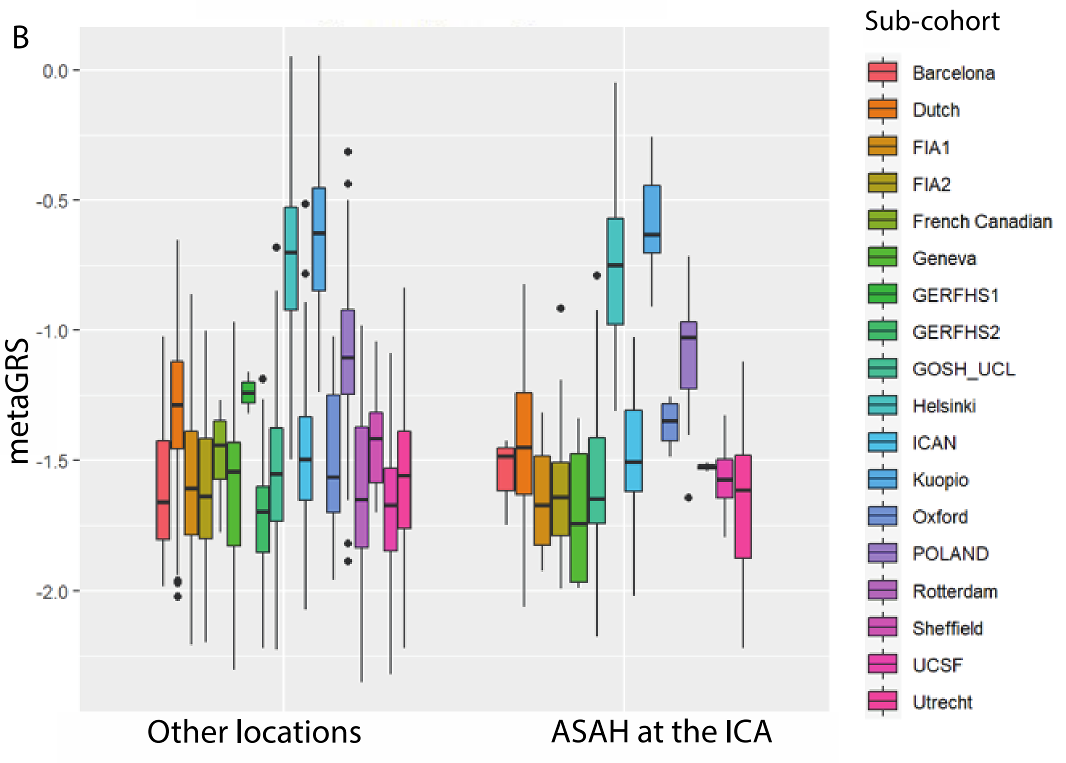

**Supplementary Figure 9. Association between metaGRS and aneurysmal subarachnoid hemorrhage (ASAH) from an intracranial aneurysm (IA) at the posterior communicating artery (PCOM) versus other locations.** A) Violin plot of the distribution of metaGRS in the phenotype cohort among persons with an ASAH at the PCOM and the remaining group. Horizontal lines denote mean and mean ± 1 standard deviation. B) Box plots showing the distribution in each sub-cohort within the phenotype cohort. Boxes contain 25th to 75th percentile and denote the median with a horizontal line. Whiskers denote smallest value greater than 1.5 times the interquartile range below the 25th percentile, and largest value smaller than 1.5 times the interquartile range above the 75th percentile.

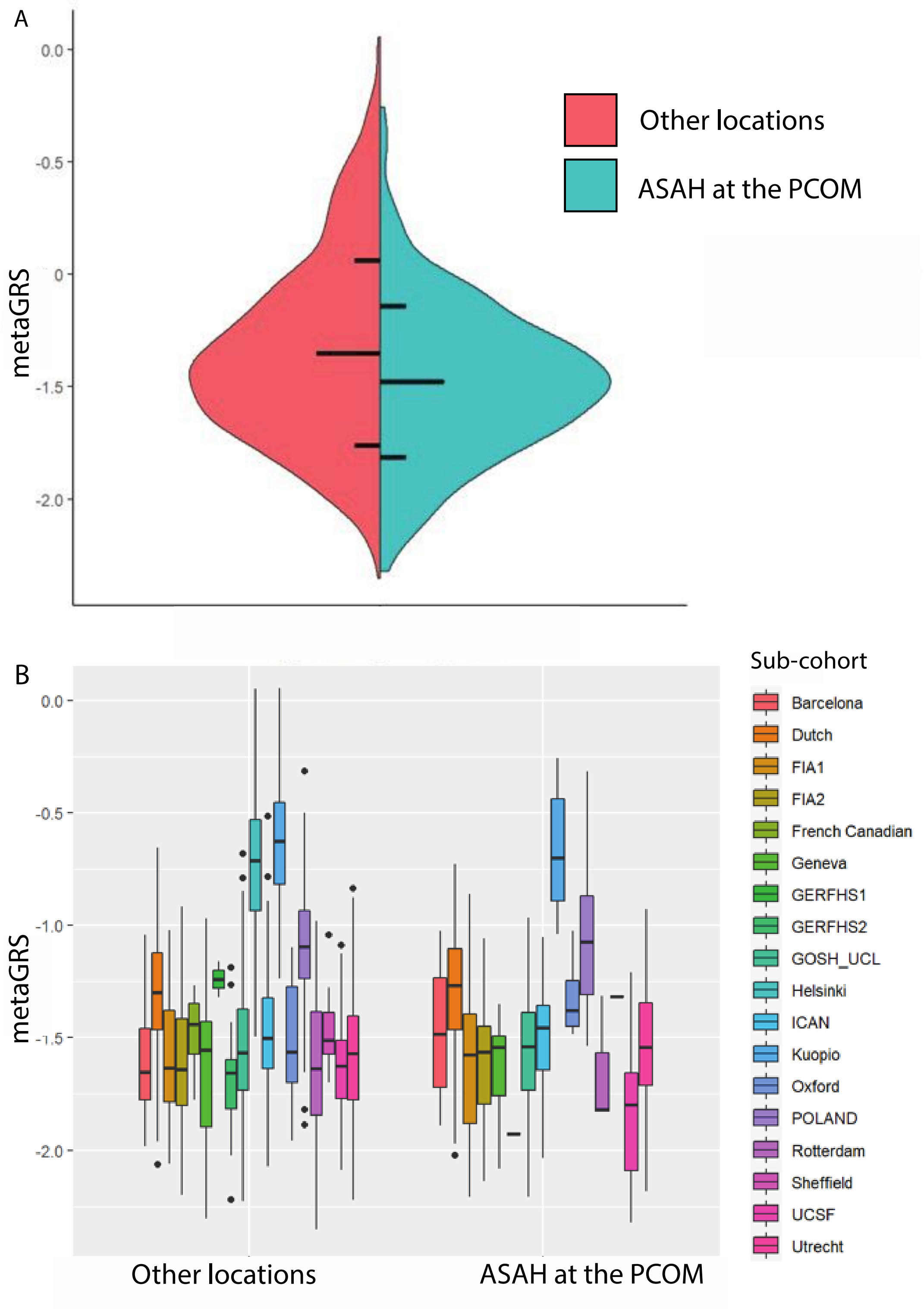

**Supplementary Figure 10. Association between metaGRS and aneurysmal subarachnoid hemorrhage (ASAH) from an intracranial aneurysm (IA) at the anterior cerebral arteries (ACA) versus other locations.** A) Violin plot of the distribution of metaGRS in the phenotype cohort among persons with an ASAH at the ACA and the remaining group. Horizontal lines denote mean and mean ± 1 standard deviation. B) Box plots showing the distribution in each sub-cohort within the phenotype cohort. Boxes contain 25th to 75th percentile and denote the median with a horizontal line. Whiskers denote smallest value greater than 1.5 times the interquartile range below the 25th percentile, and largest value smaller than 1.5 times the interquartile range above the 75th percentile.

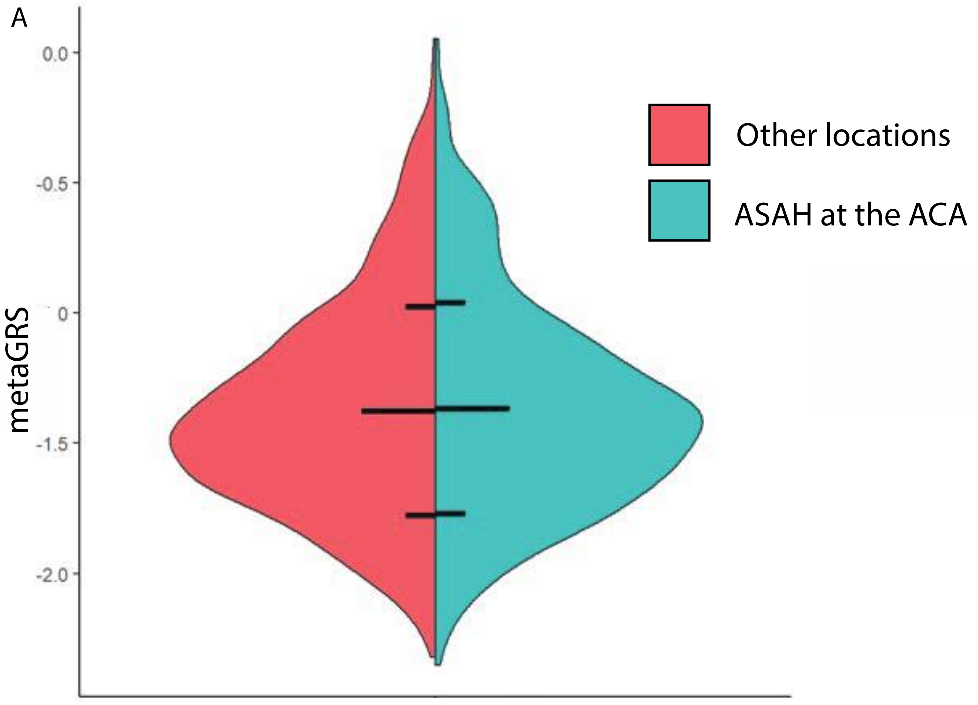

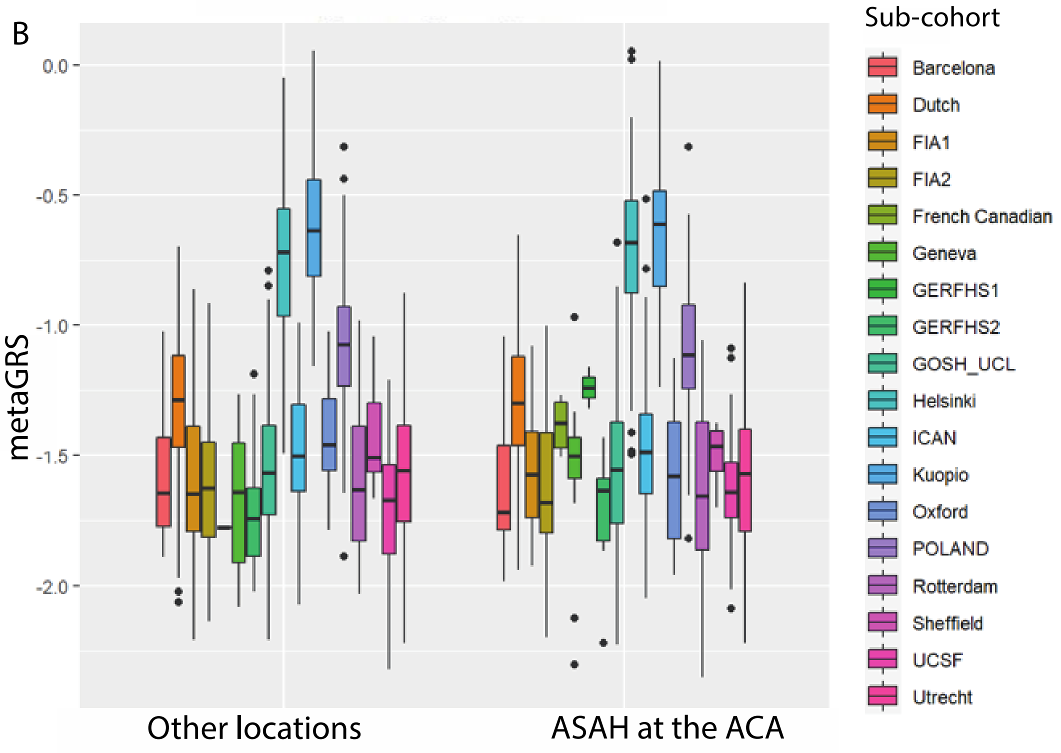

**Supplementary Figure 11. Association between metaGRS and aneurysmal subarachnoid hemorrhage (ASAH) from an intracranial aneurysm (IA) at the middle cerebral artery (MCA) versus other locations.** A) Violin plot of the distribution of metaGRS in the phenotype cohort among persons with an ASAH at the MCA and the remaining group. Horizontal lines denote mean and mean ± 1 standard deviation. B) Box plots showing the distribution in each sub-cohort within the phenotype cohort. Boxes contain 25th to 75th percentile and denote the median with a horizontal line. Whiskers denote smallest value greater than 1.5 times the interquartile range below the 25th percentile, and largest value smaller than 1.5 times the interquartile range above the 75th percentile.

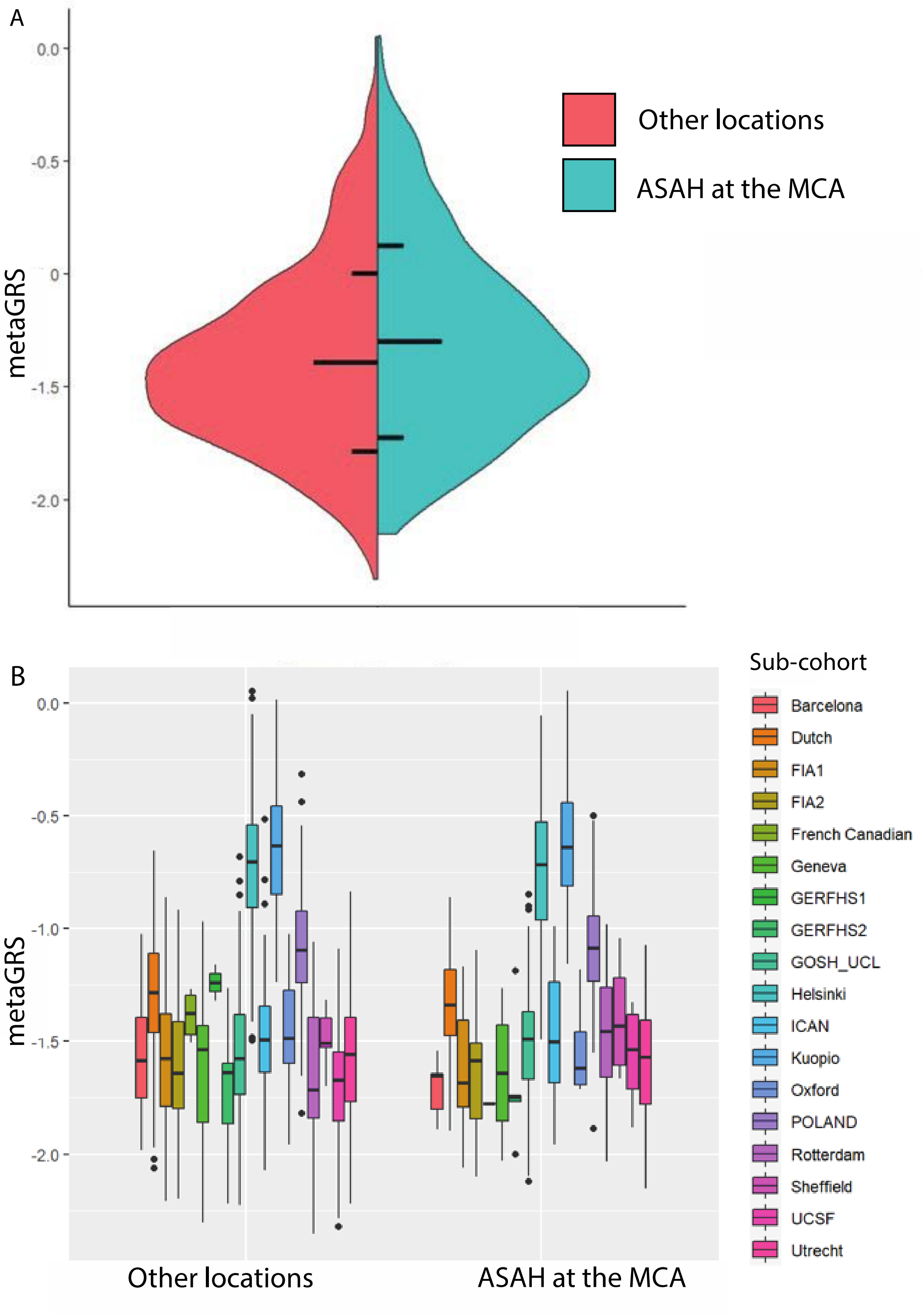

**Supplementary Figure 12. Association between metaGRS and aneurysmal subarachnoid hemorrhage (ASAH) from an intracranial aneurysm (IA) at the posterior circulation arteries (PC) versus other locations.** A) Violin plot of the distribution of metaGRS in the phenotype cohort among persons with an ASAH at the PC and the remaining group. Horizontal lines denote mean and mean ± 1 standard deviation. B) Box plots showing the distribution in each sub-cohort within the phenotype cohort. Boxes contain 25th to 75th percentile and denote the median with a horizontal line. Whiskers denote smallest value greater than 1.5 times the interquartile range below the 25th percentile, and largest value smaller than 1.5 times the interquartile range above the 75th percentile.

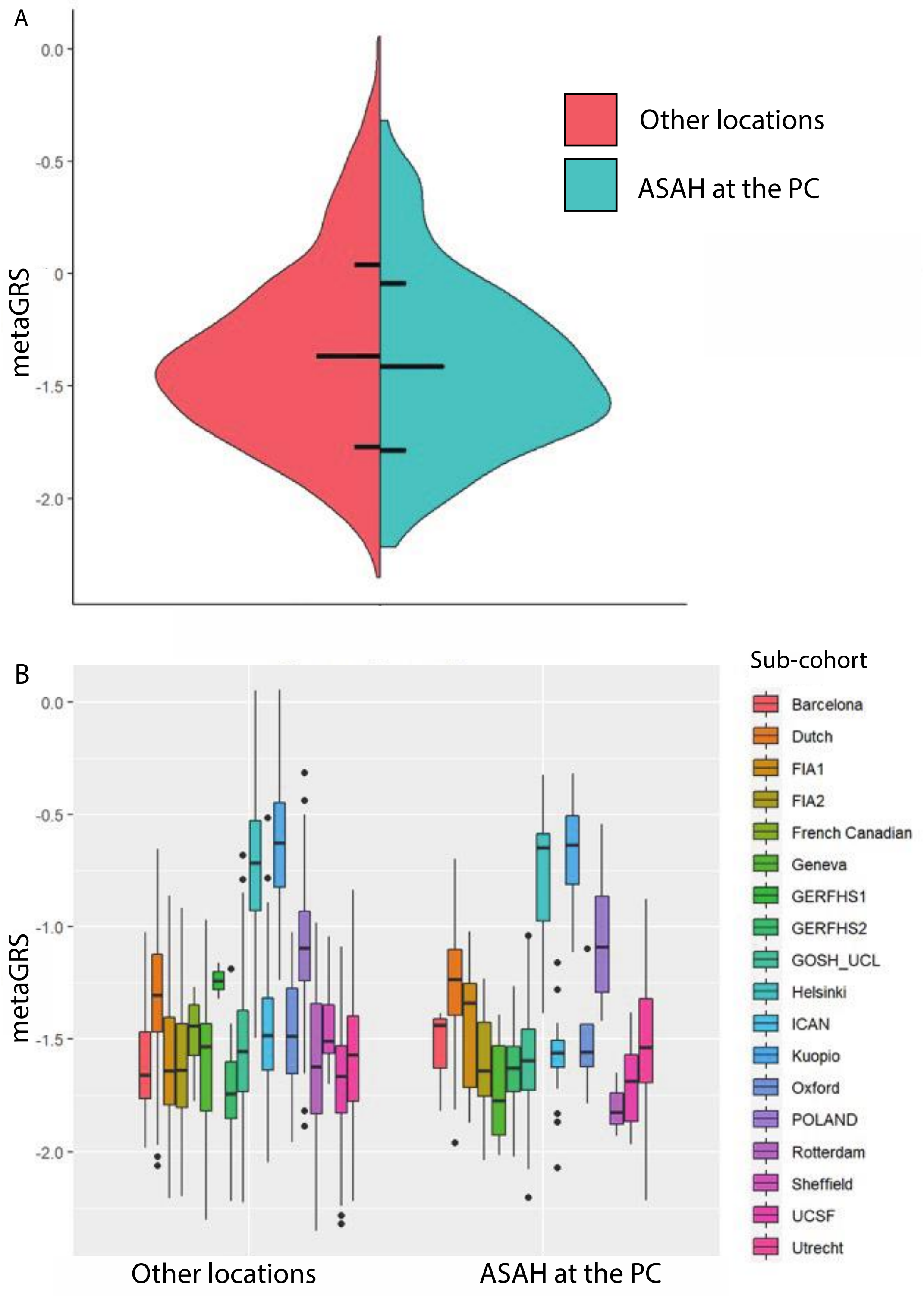

**Supplementary Figure 13. Association between metaGRS and sex among persons with an intracranial aneurysm (IA, either ruptured or unruptured).** A) Violin plot of the distribution of metaGRS in the phenotype cohort stratified by women and men. Horizontal lines denote mean and mean ± 1 standard deviation. B) Box plots showing the distribution in each sub-cohort within the phenotype cohort. Boxes contain 25th to 75th percentile and denote the median with a horizontal line. Whiskers denote smallest value greater than 1.5 times the interquartile range below the 25th percentile, and largest value smaller than 1.5 times the interquartile range above the 75th percentile.

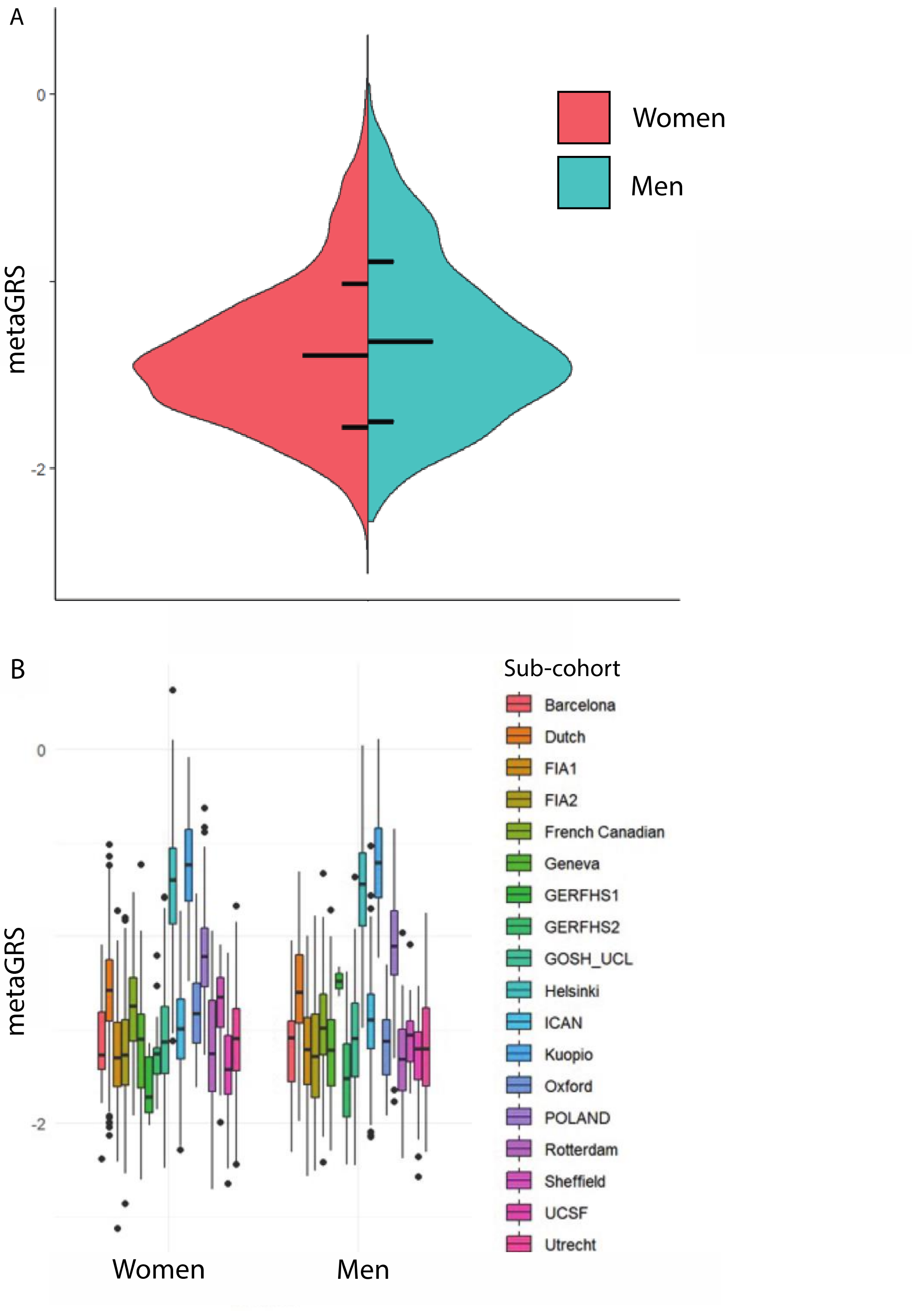

**Supplementary Figure 14. Association between metaGRS and family history among persons with an intracranial aneurysm (IA, either ruptured or unruptured).** A) Violin plot of the distribution of metaGRS in the phenotype cohort stratified by family history of IA. Horizontal lines denote mean and mean ± 1 standard deviation. B) Box plots showing the distribution in each sub-cohort within the phenotype cohort. Boxes contain 25th to 75th percentile and denote the median with a horizontal line. Whiskers denote smallest value greater than 1.5 times the interquartile range below the 25th percentile, and largest value smaller than 1.5 times the interquartile range above the 75th percentile.

A

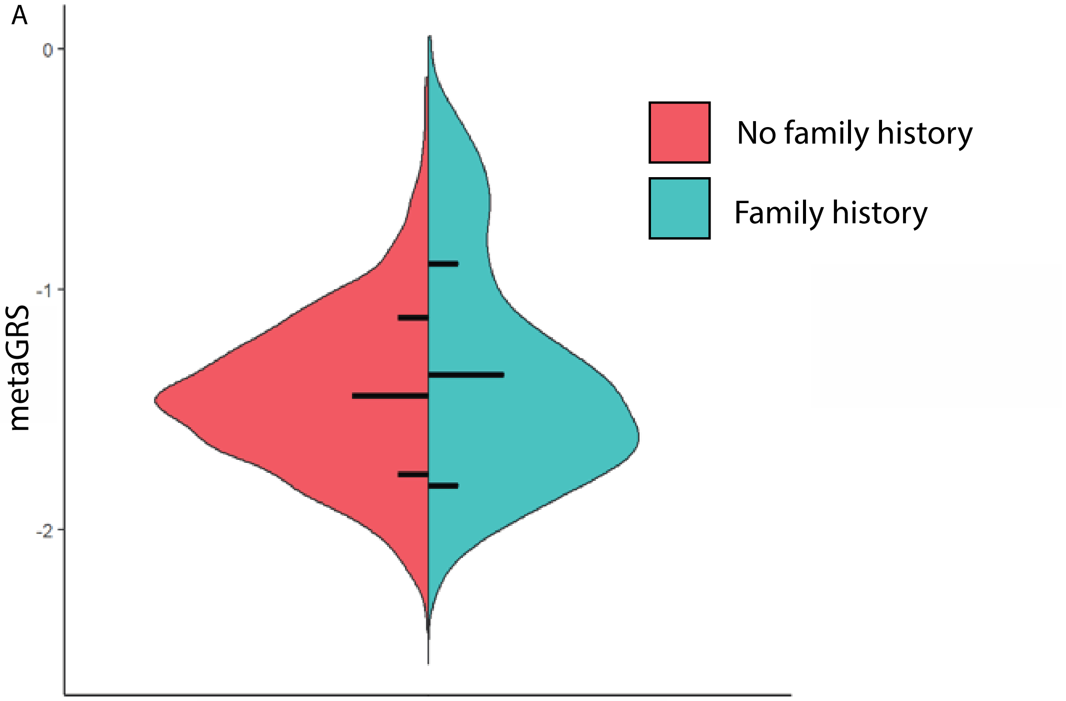

B

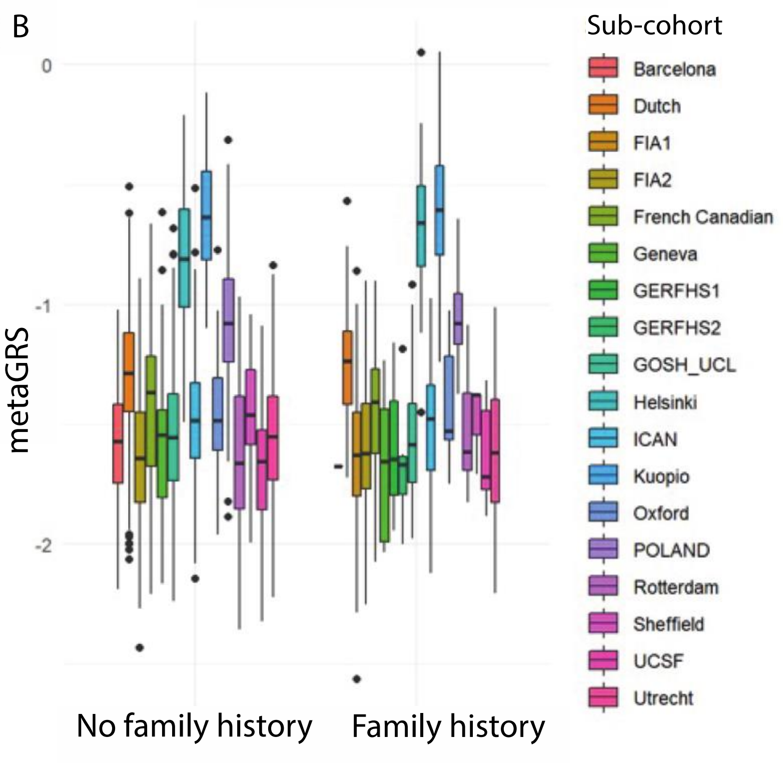

**Supplementary Figure 15. Association between metaGRS and rupture status of an intracranial aneurysm (IA).** A) Violin plot of the distribution of metaGRS in the phenotype cohort stratified by rupture status (unruptured IA, UIA versus aneurysmal subarachnoid hemorrhage, ASAH). Horizontal lines denote mean and mean ± 1 standard deviation. B) Box plots showing the distribution in each sub-cohort within the phenotype cohort. Boxes contain 25th to 75th percentile and denote the median with a horizontal line. Whiskers denote smallest value greater than 1.5 times the interquartile range below the 25th percentile, and largest value smaller than 1.5 times the interquartile range above the 75th percentile.

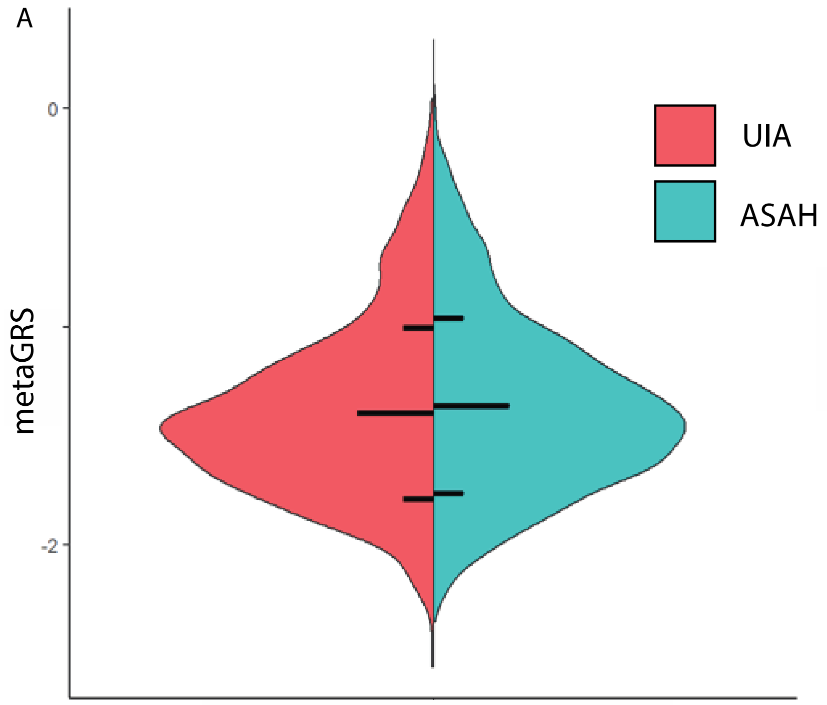

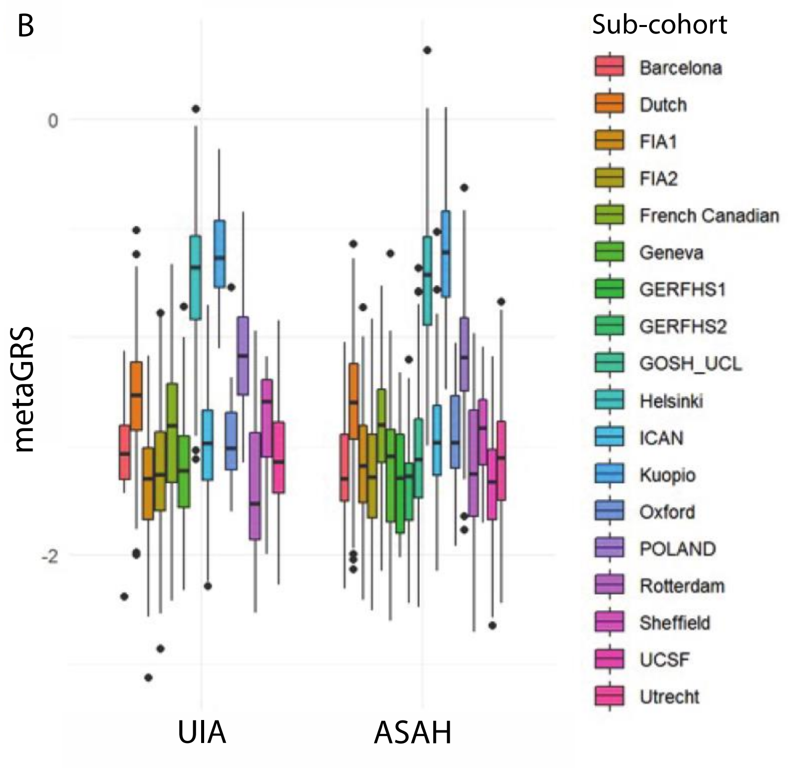

**Supplementary Figure 16. Association between metaGRS and size of intracranial aneurysm (IA) at time of aneurysmal subarachnoid hemorrhage (ASAH).** A) Darker points are (partially) overlapping data point. Line is the linear regression line of metaGRS on size at ASAH. Shaded area denotes 95% confidence interval. B) Per sub-cohort stratified linear regression of metaGRS on age at ASAH. Colored shaded area denote 95% confidence interval for each sub-cohort.

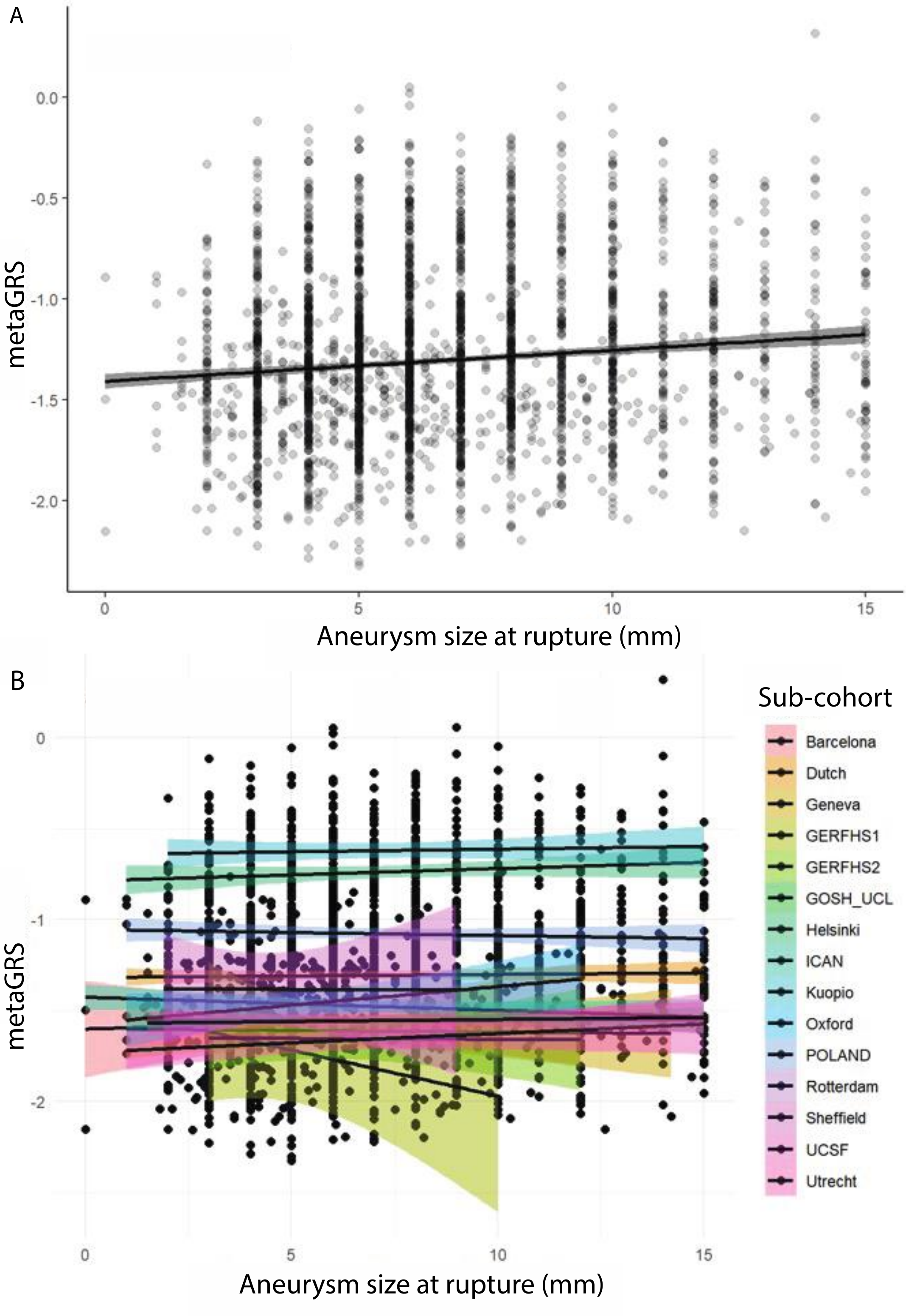

**Supplementary Figure 17. Association between metaGRS and hypertension among persons with an intracranial aneurysm (IA, either ruptured or unruptured).** A) Violin plot of the distribution of metaGRS in the phenotype cohort stratified by hypertension status. Horizontal lines denote mean and mean ± 1 standard deviation. B) Box plots showing the distribution in each sub-cohort within the phenotype cohort. Boxes contain 25th to 75th percentile and denote the median with a horizontal line. Whiskers denote smallest value greater than 1.5 times the interquartile range below the 25th percentile, and largest value smaller than 1.5 times the interquartile range above the 75th percentile.

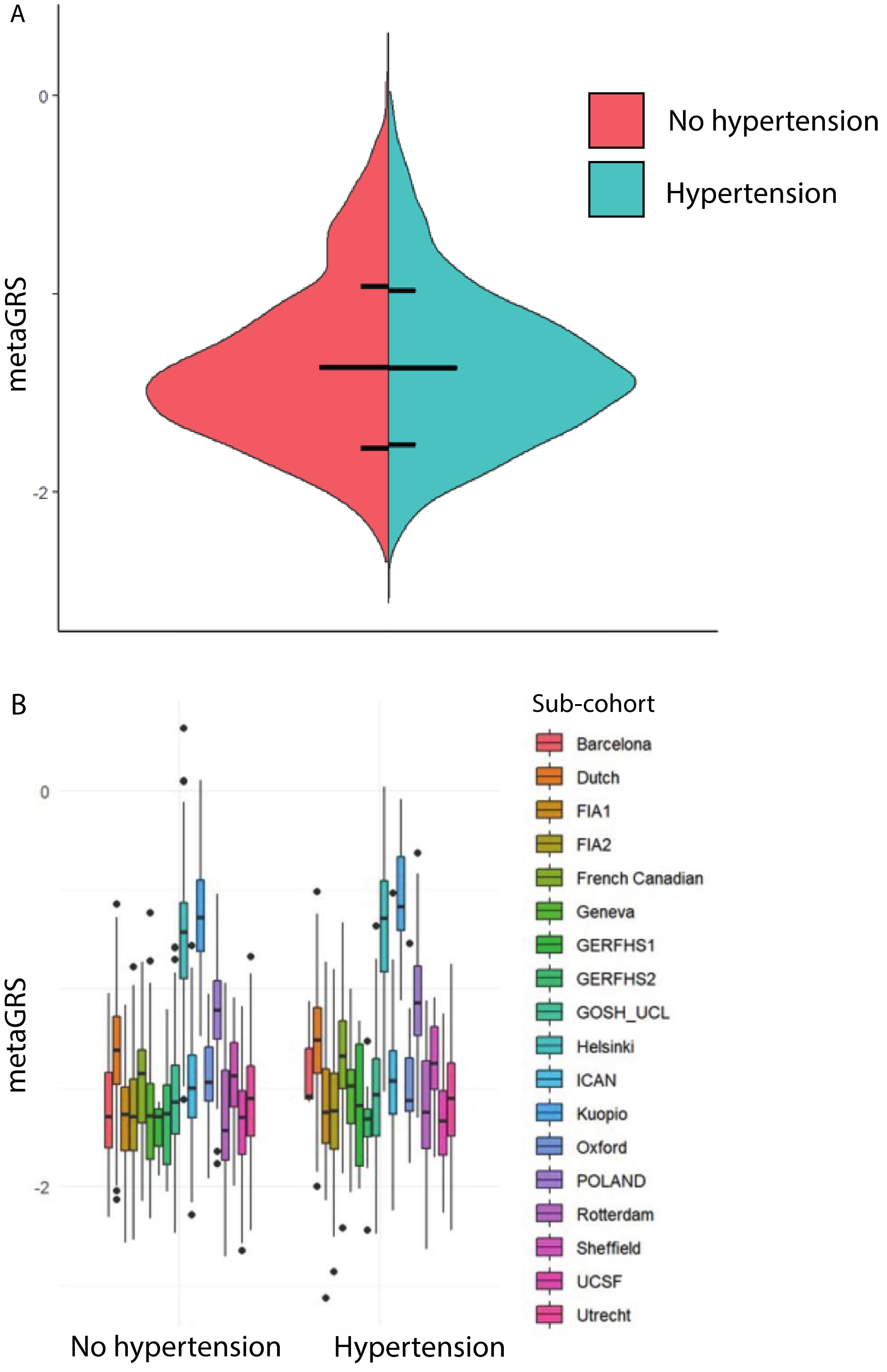

**Supplementary Figure 18. Association between metaGRS and smoking among persons with an intracranial aneurysm (IA, either ruptured or unruptured).** A) Violin plot of the distribution of metaGRS in the phenotype cohort stratified by smoking history (never, or either a current or past smoker). Horizontal lines denote mean and mean ± 1 standard deviation. B) Box plots showing the distribution in each sub-cohort within the phenotype cohort. Boxes contain 25th to 75th percentile and denote the median with a horizontal line. Whiskers denote smallest value greater than 1.5 times the interquartile range below the 25th percentile, and largest value smaller than 1.5 times the interquartile range above the 75th percentile

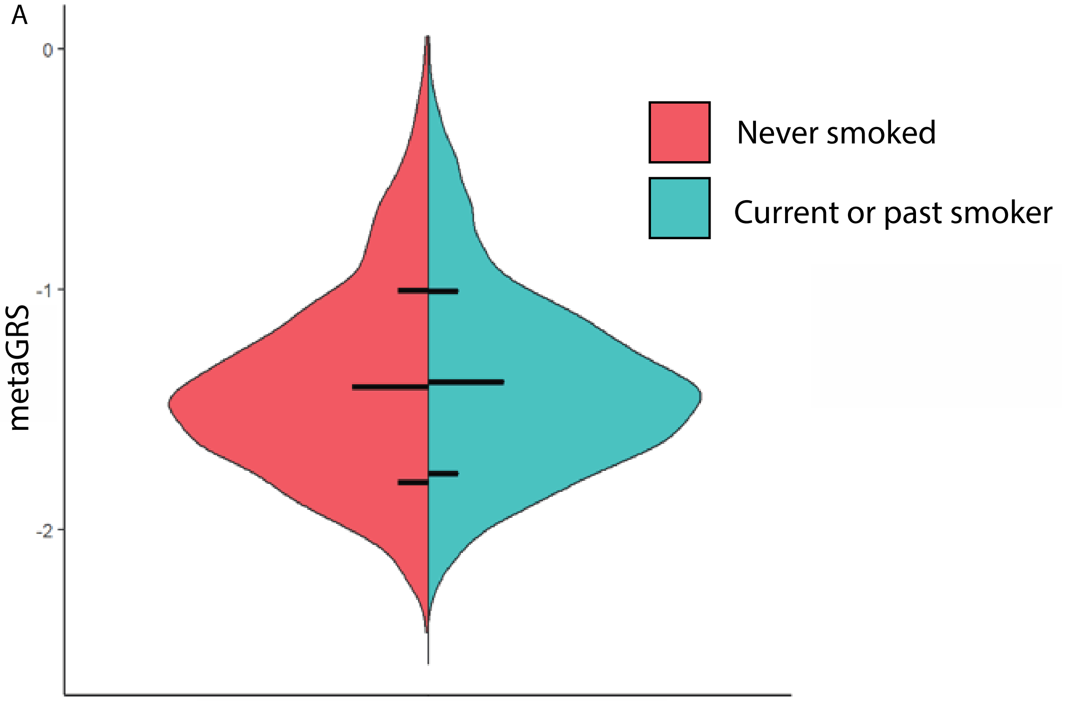

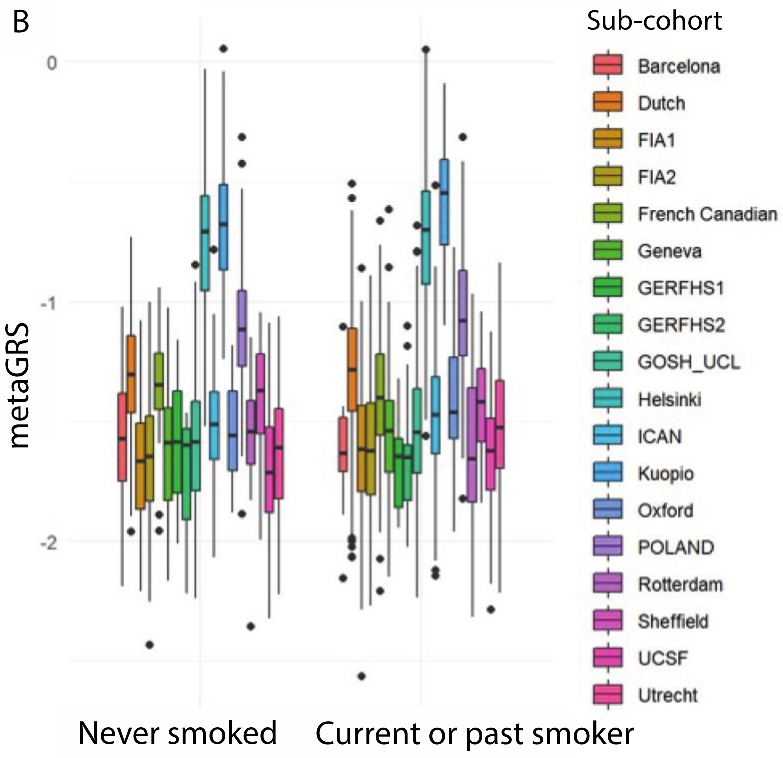

**Supplementary Figure 19. Distribution of metaGRS among sub-cohort of the phenotype cohort.** Areas are colored according to metaGRS for clarity.

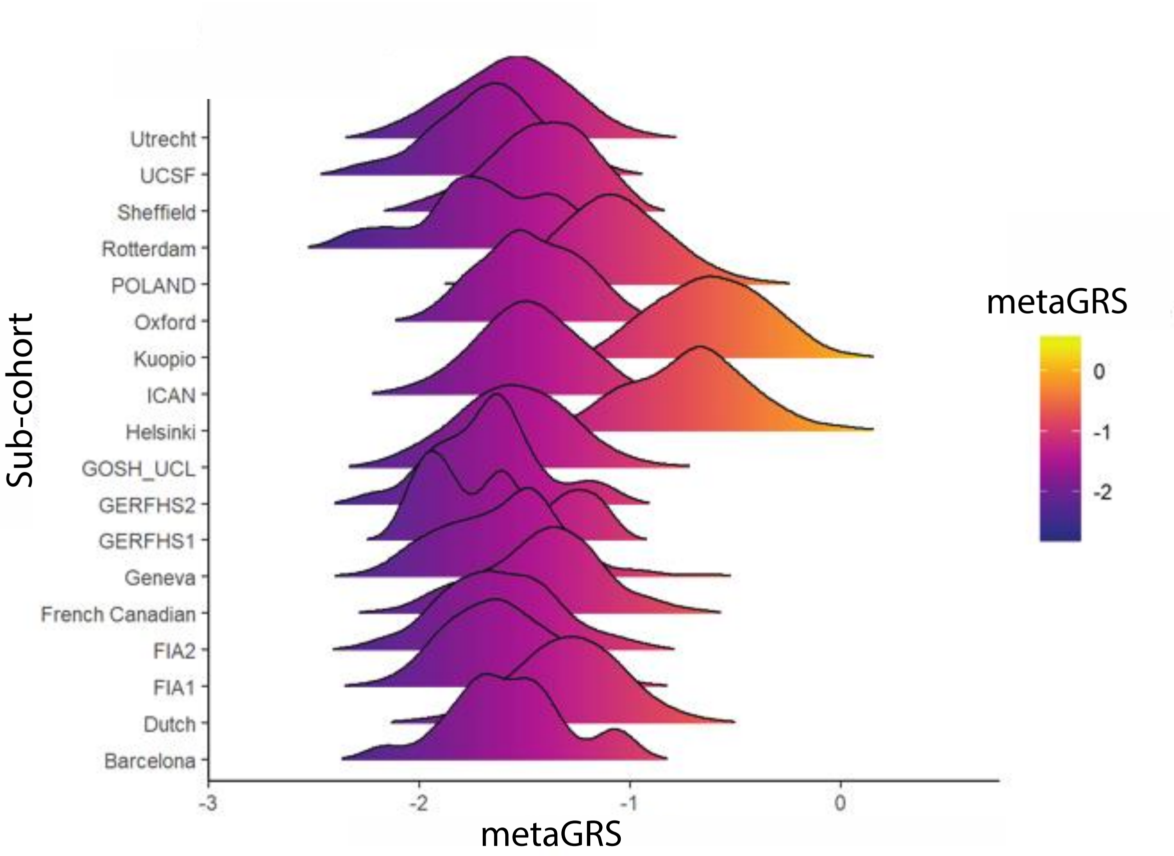
